# Spatial transcriptomics of juxtaglomerular renin cells under angiotensin receptor blockade in human IgA nephropathy

**DOI:** 10.64898/2026.11.22.26368708

**Authors:** Shaun G. Hong, Ara Ko, Myeongseop Kim, Donggun Kim, Hyunah Ku, Jae-Ik Oh, Jinsun Lee, Min Woo Kang, Youmin Shin, Jeong Ho Joo, Jeong Seok Lee, Hajeong Lee, Seung Seok Han, Yaerim Kim, Jinkyung Kwon, Jeonghwan Lee, Jung Pyo Lee, Ji In Park, Jung Tak Park, Jung Nam An, Kyung-Chul Moon, Hyun Je Kim, Young-Gon Kim, Sehoon Park, Dong Ki Kim, the KORNERSTONE investigators

## Abstract

**Background:** Renin–angiotensin system (RAS) blockade is the foundation of treatment in IgA nephropathy, yet the response of the renin cells, the macula densa and the arterioles of the human kidney to this treatment has hardly been described.

**Methods:** Single-cell-level spatial transcriptomics was obtained from archival biopsies of 154 participants, including 60 patients with IgA nephropathy, 26 of whom took an angiotensin receptor blocker. Renin cells and the macula densa were assessed at each vascular pole and along the arterioles and compared with plasma renin and the Kidney Precision Medicine Project atlas.

**Results:** Treated patients had 2.66 renin cells per glomerulus and untreated patients 0.99 (adjusted difference 1.30; 95% CI 0.42 to 2.18), with or without hypertension. The difference came from pole juxtaglomerular cells, more of which transcribed *REN* (19 percentage points; 7 to 31), with as many pole smooth muscle cells as in untreated patients. Renin cells of treated and untreated patients had a similar distribution along the arteriole and the same renin-cell program, in contrast to the changes reported in mouse renin cells. At each vascular pole, macula densa marker genes of the adjacent tubule increased with the number of renin cells, and treated patients had more renin cells without more macula densa cells. Plasma renin rose with tissue renin cells in treated and untreated patients alike, and neither was related to the composite kidney outcome.

**Conclusions:** In human IgA nephropathy, RAS blockade is accompanied by a larger population of juxtaglomerular renin cells, in which more juxtaglomerular cells transcribe renin, with the same renin-cell identity and distribution as in untreated patients, in contrast to the findings in mice. At each vascular pole, renin cells increase with macula densa marker genes of the adjacent tubule, and plasma renin rises with tissue renin cells.

## Introduction

RAS blockade is the foundation of treatment in IgA nephropathy: a maximally tolerated angiotensin receptor blocker or angiotensin-converting enzyme inhibitor is recommended for patients with proteinuria, whether or not they have hypertension [1], and trials of newer agents enroll patients already on this treatment [2]. Few patients with IgA nephropathy avoid kidney failure in their lifetime [3], so many patients take this treatment for decades. What the drug does to the renin cells that the drug stimulates, to the macula densa that controls them and to the arteriole that carries them has hardly been described in these patients.

Renin cells of the juxtaglomerular apparatus release renin when perfusion pressure falls, when less sodium chloride reaches the macula densa and when sympathetic activity rises, and angiotensin II suppresses renin release in return [4]. When this feedback is removed, renin cells increase in number, and smooth muscle cells of the afferent arteriole express renin again [5]. Angiotensin-converting enzyme inhibition recruits renin-expressing cells along the arterioles of adult rats [6]. In mice, sustained stimulation is followed by accumulation of renin-lineage cells in the arteriolar wall and concentric arteriolar thickening [7]. Long-term captopril produces the same arteriolar hypertrophy, and renin-lineage cells of mice lacking renin change their phenotype and express extracellular-matrix and secreted-protein genes [8].

In patients, the evidence comes from small series. Six patients with benign nephrosclerosis who had taken RAS inhibitors for more than five years had thicker afferent arteriolar walls and twice the renin-positive juxtaglomerular area of untreated patients [8], and juxtaglomerular hyperplasia has been described in single patients under chronic RAS blockade [9] and under dual angiotensin blockade [10]. These observations have led to the proposal that concentric arteriolar hypertrophy is a kidney risk of long-term RAS blockade [11]. Single-cell studies of RAS inhibition have been performed in a rat model of diabetic kidney disease [12], but, to our knowledge, not in human tissue.

Single-cell-level spatial transcriptomics registered to histology places each renin cell beside its glomerulus, its macula densa and its arteriole in the same section [13]. We asked how angiotensin receptor blockade before biopsy changes the number, origin and transcriptional program of juxtaglomerular renin cells in patients with IgA nephropathy, whether the macula densa takes part in this response, and whether renin cells spread along the arteriole as in mice.

## Methods

### Participants and clinical data

Participants of the spatial cohort were enrolled in the KORNERSTONE cohort [14] and underwent a kidney biopsy at Seoul National University Hospital, and single-cell-level spatial transcriptomics data were generated from archival formalin-fixed, paraffin-embedded blocks of these biopsies. The spatial cohort included 154 participants: IgA nephropathy 60, minimal change disease 25, membranous nephropathy 13, focal segmental glomerulosclerosis 4, diabetic kidney disease 28, and 24 living kidney donors with zero-time implantation biopsies as the reference. The plasma cohort included 225 patients with IgA nephropathy from KORNERSTONE hospitals with plasma collected at biopsy, and 51 of the 60 patients with IgA nephropathy in the spatial cohort were also in the plasma cohort. The study was approved by the Institutional Review Board of Seoul National University Hospital (No. 2511-060-1690), and tissue and clinical data were used based on written informed consent.

Medication before biopsy was available for 113 of the 130 patients with kidney disease, including all 60 patients with IgA nephropathy. RAS blockade was any angiotensin receptor blocker or angiotensin-converting enzyme inhibitor before biopsy, prescribed without knowledge of the tissue; all 26 treated patients with IgA nephropathy took an angiotensin receptor blocker. Hypertension, blood pressure and urine protein-to-creatinine ratio (UPCR) were recorded at biopsy, the Oxford classification [15] was taken from the SNUH IgA nephropathy registry (M, E, S and T were not available for three biopsies of 2011, when the biopsy reports did not yet give Oxford scores), and the estimated glomerular filtration rate (eGFR) at biopsy was computed from the serum creatinine at biopsy with the 2021 Chronic Kidney Disease Epidemiology Collaboration equation [16]. Plasma proteins were measured by proximity extension assay [17] as normalized protein expression (NPX) [18].

### Renin cells, macula densa and arterioles

Sections were profiled on the Xenium platform [13] with the Prime 5K panel (10x Genomics; 5,101 genes) as described [19]. Cell types were taken from the cohort annotation, which followed the cell-type scheme of the Kidney Precision Medicine Project (KPMP) [20] single-nucleus atlas [21], and in 144 participants the periodic acid–Schiff (PAS)-stained section was registered to the cell coordinates. Glomeruli were clusters of podocytes, mesangial and parietal epithelial cells (2,127 glomeruli in 152 participants), and 2,384 arterioles with an outer diameter up to 100 µm were outlined on PAS by automated segmentation (Supplementary Methods).

One of the 24 cell types of the cohort annotation, the juxtaglomerular label, marked juxtaglomerular cells by their expression profile (Supplementary Methods). A renin cell was a cell with this label and at least one nuclear *REN* transcript; at least three transcripts, with or without the label, served as a stricter definition. Renin cells, labeled cells without a *REN* transcript and smooth muscle cells within 20 µm of a glomerulus were the cells of the vascular pole. Renin cells were also counted per mm^2^ of tissue by distance from the nearest glomerulus and in the wall of each arteriole; arterioles more than 20 µm from every glomerulus with at least three renin cells in the wall were called distant renin arterioles.

The macula densa was defined by position, without marker genes: thick ascending limb cells within 15 µm of a glomerulus and within 40 µm of its vascular pole, located from the smooth muscle cells and labeled cells without a *REN* transcript. Its genes were obtained from the whole panel against cortical thick ascending limb cells.

### Expression analyses and external data

Nuclear transcripts were summed per participant and cell population and compared with limma [22] and voom weights [23]; gene sets of the Gene Ontology, Reactome and Hallmark collections [24] were tested on the ranked genes. Each renin cell was placed on a score from arteriolar smooth muscle (0) to the average renin cell (1). At each vascular pole, thick ascending limb cells within 15 µm of the glomerulus were related to the number of renin cells at the same pole. Ligand–receptor pairs were taken from CellChatDB v2 [25]. In the KPMP atlas, renin nuclei were compared by the open-access field for RAS blockade (69 participants on treatment, 47 not). Mouse renin-cell gene sets after renin deletion [8], after captopril with a low-salt diet for two weeks [26] or one week [27] and after lowered perfusion pressure [28] were mapped to human orthologs.

### Statistical analysis

Participant measures were compared by least squares with heteroskedasticity-consistent errors, adjusted for age, sex and sequencing batch (basic model) and additionally for mean arterial pressure, log UPCR and eGFR (adjusted model); renin-cell counts were also modeled by negative binomial regression with the number of glomeruli as exposure. Analyses of non-diabetic glomerular disease included a diagnosis term, and living donors served as a descriptive reference. The kidney outcome was a composite of an eGFR decline of 40% or more from the eGFR at biopsy, confirmed at least 28 days later or, at the end of follow-up, without recovery to 80%, or an eGFR below 15 mL/min/1.73 m^2^ on two occasions (Supplementary Methods), analyzed by Cox models. In the spatial cohort, juxtaglomerular measures and plasma renin were related, per standard deviation, to the clinical state at biopsy and the kidney course, adjusted for age, sex, batch, RAS blockade, eGFR and log UPCR at biopsy (Supplementary Methods). False discovery rates were controlled by the Benjamini–Hochberg method [29].

## Results

### Patients

Of the 60 patients with IgA nephropathy, 26 took an angiotensin receptor blocker before biopsy, 9 without and 17 with hypertension; 4 had hypertension without RAS blockade and 30 had neither (Table 1). Treated patients were older and had a lower eGFR, with similar UPCR. Of the 34 untreated patients, 27 started RAS blockade after biopsy. Plasma renin was higher in treated patients, by 0.86 NPX in the spatial cohort (95% CI 0.38 to 1.34) and by 0.58 NPX in the plasma cohort (0.30 to 0.86).

**Table 1.** Characteristics of the participants.

|  |  |  |  |  |  |  |  |  |
|---|---|---|---|---|---|---|---|---|
| *A. IgA nephropathy of the spatial cohort by treatment group* |  |  |  |  |  |  |  |  |
|  |  | RAS blockade |  |  | No RAS blockade |  |  |  |
| Characteristic | All patients | All | No hypertension | Hypertension | All | No hypertension | Hypertension |  |
| Patients, *n* | 60 | 26 | 9 | 17 | 34 | 30 | 4 |  |
| Age, years | 39.6 (33.8, 50.1) | 43.8 (37.2, 54.2) | 41.0 (29.0, 50.0) | 45.9 (41.9, 54.2) | 36.2 (30.5, 43.8) | 35.9 (28.8, 41.9) | 51.3 (47.0, 53.4) |  |
| Female, *n* (%) | 30 (50) | 10 (38) | 4 (44) | 6 (35) | 20 (59) | 18 (60) | 2 (50) |  |
| eGFR, mL/min/1.73 m<sup>2</sup> | 93.2 (79.2, 113.0) | 81.9 (68.0, 93.0) | 91.1 (82.1, 102.9) | 77.2 (61.7, 82.4) | 109.8 (89.9, 122.5) | 112.1 (95.2, 123.2) | 82.3 (69.2, 93.7) |  |
| UPCR, g/g | 0.86 (0.49, 1.19) | 0.91 (0.61, 0.99) | 0.66 (0.56, 0.93) | 0.93 (0.75, 3.40) | 0.77 (0.38, 1.56) | 0.64 (0.35, 1.56) | 0.96 (0.94, 1.23) |  |
| MAP, mmHg | 86.3 (78.6, 93.8) | 85.0 (78.8, 93.9) | 81.0 (78.3, 84.3) | 88.0 (82.3, 95.0) | 86.7 (78.4, 92.5) | 86.5 (78.4, 89.2) | 96.2 (89.4, 100.8) |  |
| CCB, *n* | 11 | 9 | 0 | 9 | 2 | 0 | 2 |  |
| Loop/thiazide diuretic, *n* | 3 | 2 | 0 | 2 | 1 | 0 | 1 |  |
| Oxford classification, *n*/*N* |  |  |  |  |  |  |  |  |
| M1 | 14/57 | 6/26 | 4/9 | 2/17 | 8/31 | 7/27 | 1/4 |  |
| E1 | 11/57 | 7/26 | 2/9 | 5/17 | 4/31 | 3/27 | 1/4 |  |
| S1 | 38/57 | 20/26 | 7/9 | 13/17 | 18/31 | 14/27 | 4/4 |  |
| T1–2 | 6/57 | 3/26 | 1/9 | 2/17 | 3/31 | 3/27 | 0/4 |  |
| C1–2 | 9/60 | 3/26 | 1/9 | 2/17 | 6/34 | 6/30 | 0/4 |  |
| Plasma renin, NPX | 0.45 (−0.25, 1.03) | 0.80 (0.55, 1.67) | 0.72 (0.24, 1.03) | 1.06 (0.58, 1.78) | −0.17 (−0.35, 0.40) | −0.01 (−0.30, 0.52) | −0.61 (−0.85, −0.28) |  |
| Measured, *n* | 51 | 25 | 9 | 16 | 26 | 22 | 4 |  |
| Glomeruli per patient | 12.0 (7.5, 17.5) | 10.0 (7.0, 13.0) | 13.0 (10.0, 17.0) | 8.0 (5.0, 13.0) | 13.0 (11.0, 19.0) | 14.0 (11.0, 19.8) | 8.0 (6.5, 13.5) |  |
| Arterioles per patient | 16.0 (9.0, 30.0) | 16.5 (10.0, 42.5) | 15.0 (13.0, 26.0) | 18.0 (9.5, 47.0) | 16.0 (7.5, 27.0) | 15.0 (6.8, 26.5) | 24.0 (18.5, 32.5) |  |
| *B. Other participants of the spatial cohort and the plasma cohort* |  |  |  |  |  |  |  |  |
|  |  |  |  | eGFR, |  | RAS blockade, | Glomeruli | Arterioles |
| Group | *n* | Age, years | Female, *n* (%) | mL/min/1.73 m<sup>2</sup> | UPCR, g/g | *n*/*N* | per participant | per participant |
| MCD | 25 | 41.8 (31.4, 57.0) | 9 (36) | 103.5 (81.2, 115.5) | 8.55 (5.80, 11.63) | 7/19 | 14.0 (10.0, 19.0) | 10.0 (5.0, 14.0) |
| MN | 13 | 59.7 (53.0, 65.3) | 4 (31) | 102.9 (97.2, 106.3) | 4.78 (4.07, 7.07) | 6/10 | 11.0 (6.0, 16.0) | 9.0 (5.8, 13.5) |
| FSGS | 4 | 31.9 (20.3, 44.9) | 4 (100) | 78.9 (48.1, 114.0) | 4.35 (1.73, 8.10) | 1/1 | 14.0 (12.2, 17.0) | 15.0 (14.2, 16.0) |
| DKD | 28 | 51.3 (38.4, 59.8) | 8 (29) | 54.1 (39.6, 84.2) | 3.99 (3.04, 8.21) | 16/23 | 9.0 (5.0, 14.5) | 10.0 (7.0, 22.8) |
| Living donors | 24 | 55.0 (50.0, 58.2) | 17 (71) | 102.9 (98.8, 106.0) | – | – | 21.0 (12.5, 27.2) | 15.0 (9.0, 19.5) |
| IgAN, plasma cohort | 225 | 43.0 (35.0, 52.0) | 118 (52) | 82.2 (57.6, 104.8) | 1.18 (0.56, 2.24) | 109/225 | – | – |
| IgAN, outcome analysis | 149 | 43.0 (33.0, 51.0) | 79 (53) | 82.4 (60.6, 105.3) | 1.18 (0.66, 2.57) | 74/149 | – | – |

Values are median (lower quartile, upper quartile) or counts. All treated patients took an angiotensin receptor blocker. Hypertension, the recorded history at biopsy. Oxford classification (MEST-C) from the SNUH IgA nephropathy registry (M, E, S and T not available for three biopsies of 2011); *N*, patients with the score recorded. Plasma renin by proximity extension assay within 7 days of biopsy. Glomeruli, glomeruli of the analyzed section (one untreated patient with IgA nephropathy had none); arterioles, arterioles with an outer diameter up to 100 µm, in patients with a section registered to PAS histology. B, living donors: age and sex at donation, eGFR from pre-donation creatinine; plasma cohort: patients with IgA nephropathy with plasma within 7 days of biopsy and a drug record, 51 of them also in the spatial cohort; outcome analysis: patients of the plasma cohort followed at Seoul National University Hospital (Supplementary Methods); –, not available. CCB, calcium-channel blocker; DKD, diabetic kidney disease; eGFR, estimated glomerular filtration rate (CKD-EPI 2021); FSGS, focal segmental glomerulosclerosis; IgAN, IgA nephropathy; MAP, mean arterial pressure; MCD, minimal change disease; MN, membranous nephropathy; NPX, normalized protein expression; PAS, periodic acid–Schiff; RAS, renin–angiotensin system; UPCR, urine protein-to-creatinine ratio.

### More renin cells at the vascular pole with RAS blockade

Renin cells lay at the glomerular vascular pole between the macula densa and the arteriole (Figure 1; Figure 2a). Treated patients had 2.66 renin cells per glomerulus, untreated patients 0.99 and living donors 0.85, an adjusted difference of 1.30 (0.42 to 2.18; rate ratio 2.40; 1.55 to 3.70) and of 1.34 (0.28 to 2.40) with blood pressure, proteinuria and eGFR also in the model (Supplementary Table S1). Treated patients had more renin cells with or without hypertension (2.42 and 3.12 per glomerulus), whereas patients with hypertension who were not on RAS blockade had 0.64 (three with a glomerulus in the section; Figure 2b).

**Figure 1.**
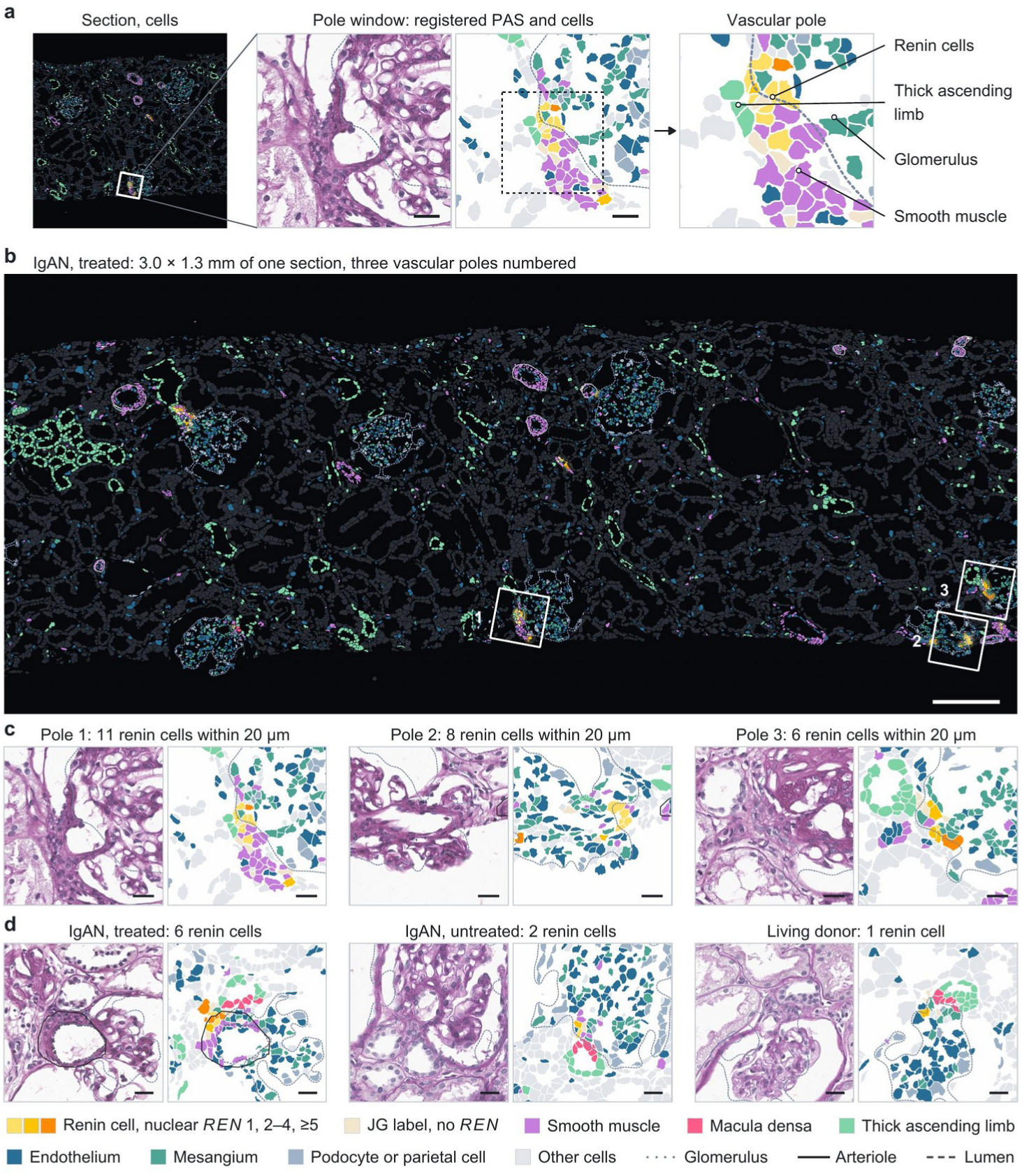
Renin cells at the glomerular vascular pole. **a**, Study design: a Xenium section registered to periodic acid–Schiff (PAS) histology, the vascular pole of one glomerulus, and the cells and structures measured there. **b**, A 3.0 × 1.3 mm window of a section from a treated patient with IgA nephropathy, every segmented cell filled by identity; numbered boxes, the poles enlarged in **c**. **c**, The three vascular poles of **b**: PAS (left) and segmented cells (right); title, renin cells within 20 µm of the glomerulus. **d**, A representative vascular pole of a treated patient, an untreated patient and a living donor; title, renin cells at the pole. Colors as in the key. Bars, 200 µm (**b**) and 20 µm (**a**, **c**, **d**). IgAN, IgA nephropathy; JG, juxtaglomerular.

**Figure 2.**
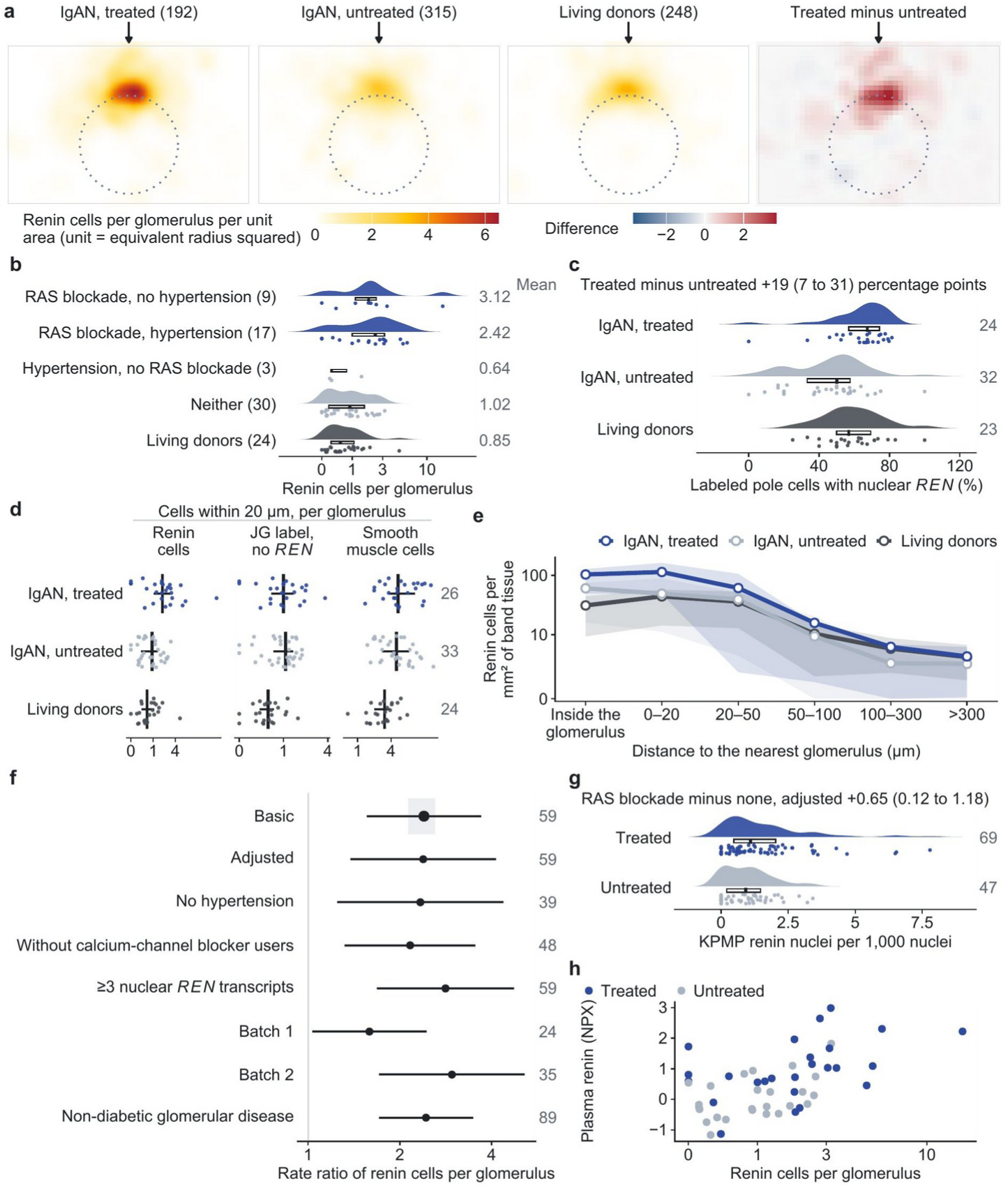
More renin cells at the vascular pole with RAS blockade. **a**, Aggregate vascular pole: renin cells around glomeruli scaled to their equivalent radius and rotated so that the vascular pole points up (arrow), by group, and the treated-minus-untreated map; glomeruli in parentheses. **b**, Renin cells per glomerulus in patients with IgA nephropathy on RAS blockade without and with hypertension, with hypertension without RAS blockade, with neither, and in living donors; participants in parentheses, means at right. **c**, Share of juxtaglomerular-labeled pole cells with a nuclear *REN* transcript; participants at right. **d**, Mural cells of the vascular pole per glomerulus by group (participants with median and interquartile range; participants at right); 33 of the 34 untreated patients had a glomerulus in the section. **e**, Renin cells per mm^2^ of tissue by distance from the glomerular boundary; lines, group means; bands, interquartile ranges of participants. **f**, Treated-to-untreated rate ratio of renin cells per glomerulus under other models, definitions, sequencing batches and patient groups (count model, logarithmic axis; shaded, the basic rate ratio with each patient left out in turn; patients at right; differences by least squares in Supplementary Table S4). **g**, Kidney Precision Medicine Project: renin nuclei per 1,000 nuclei by RAS blockade; participants at right. **h**, Plasma renin against renin cells per glomerulus in the 50 patients with IgA nephropathy with a glomerulus in the section and plasma, by treatment. Rainclouds (**b**, **c**, **g**): density, interquartile box with median, and participants (points). Points and lines (**f**), estimates and 95% confidence intervals. IgAN, IgA nephropathy; JG, juxtaglomerular; KPMP, Kidney Precision Medicine Project; NPX, normalized protein expression; RAS, renin–angiotensin system.

The difference came from juxtaglomerular cells that transcribed renin. Juxtaglomerular-labeled cells at the pole were 3.80 per glomerulus in treated and 2.00 in untreated patients (difference 1.26; 0.10 to 2.42), and 19 percentage points more of them carried a nuclear *REN* transcript in treated patients (7 to 31; Figure 2c), whereas labeled cells without a *REN* transcript were as many (1.14 and 1.01). Pole smooth muscle cells did not differ (0.02 per glomerulus; −1.61 to 1.65; Figure 2d). Among the other named cell classes of the pole, only glomerular mesangial cells were more numerous in treated patients (2.9 per pole; 0.3 to 5.6), but not in the adjusted model (Supplementary Figure S1). Outside the glomerulus, renin-cell density fell with distance in every group (Figure 2e). The difference was in the same direction in both sequencing batches, held with the stricter renin-cell definition, in non-diabetic glomerular disease (difference 1.37; 0.64 to 2.09) and among patients not taking a calcium-channel blocker (rate ratio 2.16; 1.31 to 3.55; Figure 2f; Supplementary Figure S2; Supplementary Tables S2–S4). In the KPMP atlas, participants on RAS blockade had 0.65 more renin nuclei per 1,000 nuclei (0.12 to 1.18; 116 participants; Figure 2g; Supplementary Figure S3).

Plasma renin rose with the number of renin cells in the tissue (Figure 2h). The correlation was similar in treated (Spearman ρ 0.47) and untreated patients with IgA nephropathy (ρ 0.40; 25 each with a glomerulus in the section and plasma), and the relation held with RAS blockade in the model (1.23 renin cells per glomerulus per NPX; 0.13 to 2.34). Plasma renin was not related to macula densa cells, pole smooth muscle cells or distant renin arterioles (Supplementary Table S4).

### The same renin-cell identity in treated and untreated patients

Against arteriolar smooth muscle of the same patients, renin cells expressed *REN*, *TRPM3*, the potassium channel subunits *KCNJ2* and *ABCC8*, the adenosine A1 receptor *ADORA1*, connexin 40 (*GJA5*), *MECOM* and *GATA3*, and had less *ACTA2*, *VIM*, *JAG1* and *COL1A1* (Figure 3a; Supplementary Table S5). The profile matched renin nuclei against vascular smooth muscle nuclei in KPMP (Spearman ρ 0.70 over 335 genes; Figure 3b).

**Figure 3.**
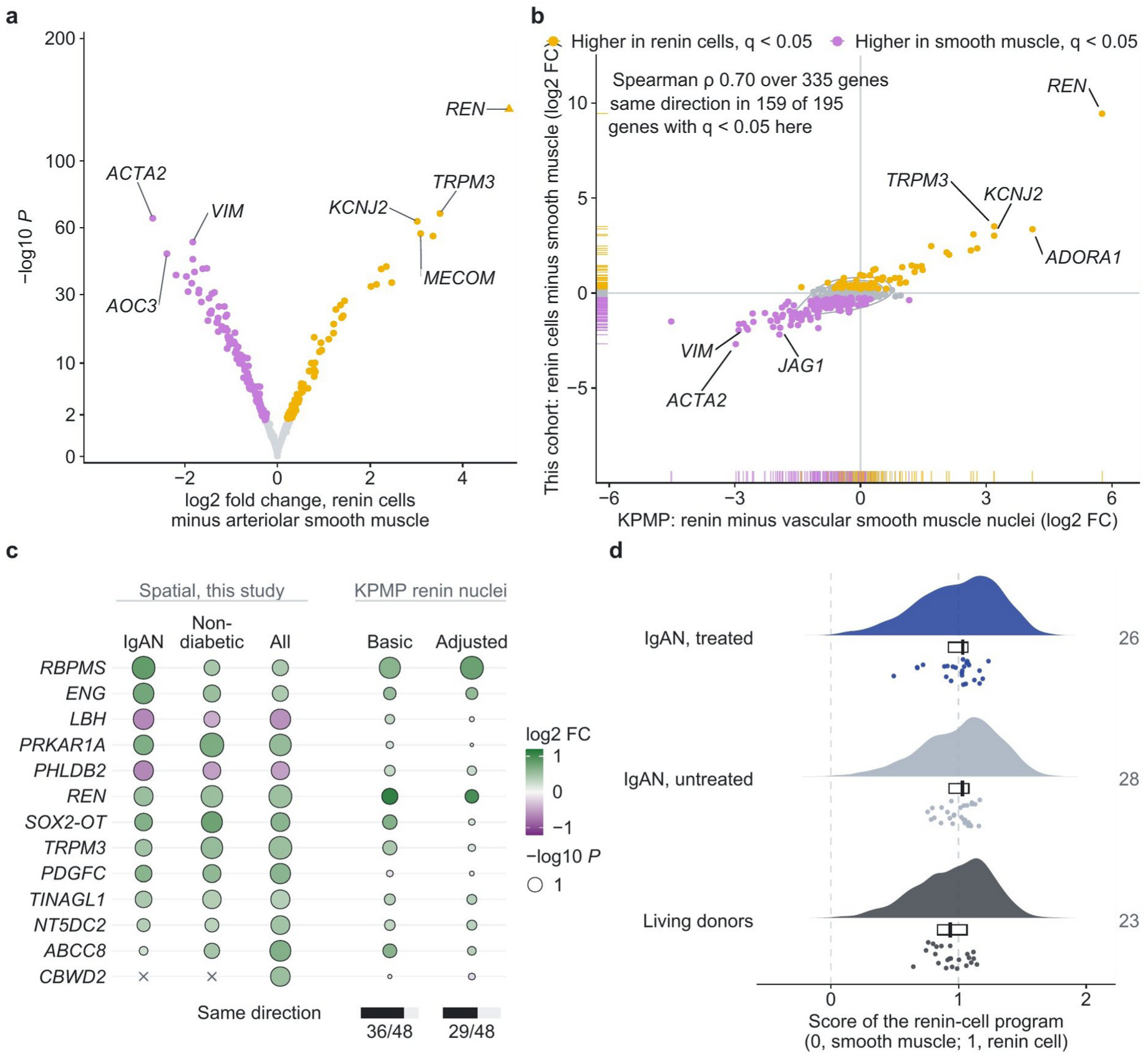
Renin-cell program. **a**, Renin cells against arteriolar smooth muscle of the same patients; colors as in **b**; triangle, beyond the axis. **b**, The same differences against Kidney Precision Medicine Project (KPMP) renin minus vascular smooth muscle nuclei. **c**, Renin cells of treated against untreated patients in IgA nephropathy, non-diabetic glomerular disease and all diagnoses, and the direction in KPMP renin nuclei: the genes with the ten smallest *P* values across diagnoses and the six smallest in IgA nephropathy, in the order of *P* in IgA nephropathy; color, log_2_ fold change; size, −log_10_ *P*; ×, not tested; below, the genes in the same direction in KPMP among the 48 measured there. **d**, Cell-level score of the renin-cell program (0, average arteriolar smooth muscle cell; 1, average renin cell) of renin cells by group: density, interquartile box with median, and participant medians (points); participants at right. FC, fold change; IgAN, IgA nephropathy.

In treated patients, the genes with the smallest *P* values in renin cells were *RBPMS*, *ENG* and *PRKAR1A*, the regulatory subunit of protein kinase A, with lower *LBH* and *PHLDB2*, and *REN* was higher (log_2_ fold change 0.52; 0.09 to 0.95); none of the 145 genes tested in IgA nephropathy or of the 139 tested in non-diabetic glomerular disease reached a false discovery rate of 10% (Figure 3c; Supplementary Table S6). *RBPMS* and *REN* were also higher in KPMP renin nuclei of participants on RAS blockade, and 36 of the 48 genes with the smallest *P* values that were measured there changed in the same direction (29 with adjustment). The mouse renin-cell profiles after renin deletion, after captopril with a low-salt diet (two data sets) and after low perfusion pressure were not related to the differences by treatment (Spearman ρ −0.05 to 0.09), and neither the genes higher after two weeks of captopril (12 testable) nor the matrix-secretory genes (13) were enriched. The 5 secreted-protein genes of the matrix-secretory set were higher as a set (*P* 0.04; adjusted 0.04), four of the five in that direction, *BGN* among them (log_2_ fold change 0.57; 0.09 to 1.05; Supplementary Figure S4; Supplementary Table S7); *BGN* was in the same direction in KPMP renin nuclei of participants on RAS blockade (0.64; *P* 0.05).

On a cell-level score of the renin-cell program, renin cells of treated and untreated patients were the same (difference 0.02; −0.08 to 0.11; Figure 3d). No gene set differed by treatment at a false discovery rate of 10%.

### Macula densa and renin cells at the same vascular pole

Thick ascending limb cells at the vascular pole had higher *ROBO2*, *MEST*, *CALCR* and *STIM2* and lower *UMOD* and *EGF* than cortical thick ascending limb cells (49 participants; Figure 4a; Supplementary Table S8). The macula densa lay a median of 16 µm from the nearest renin cell (Figure 4b; Supplementary Figure S5).

**Figure 4.**
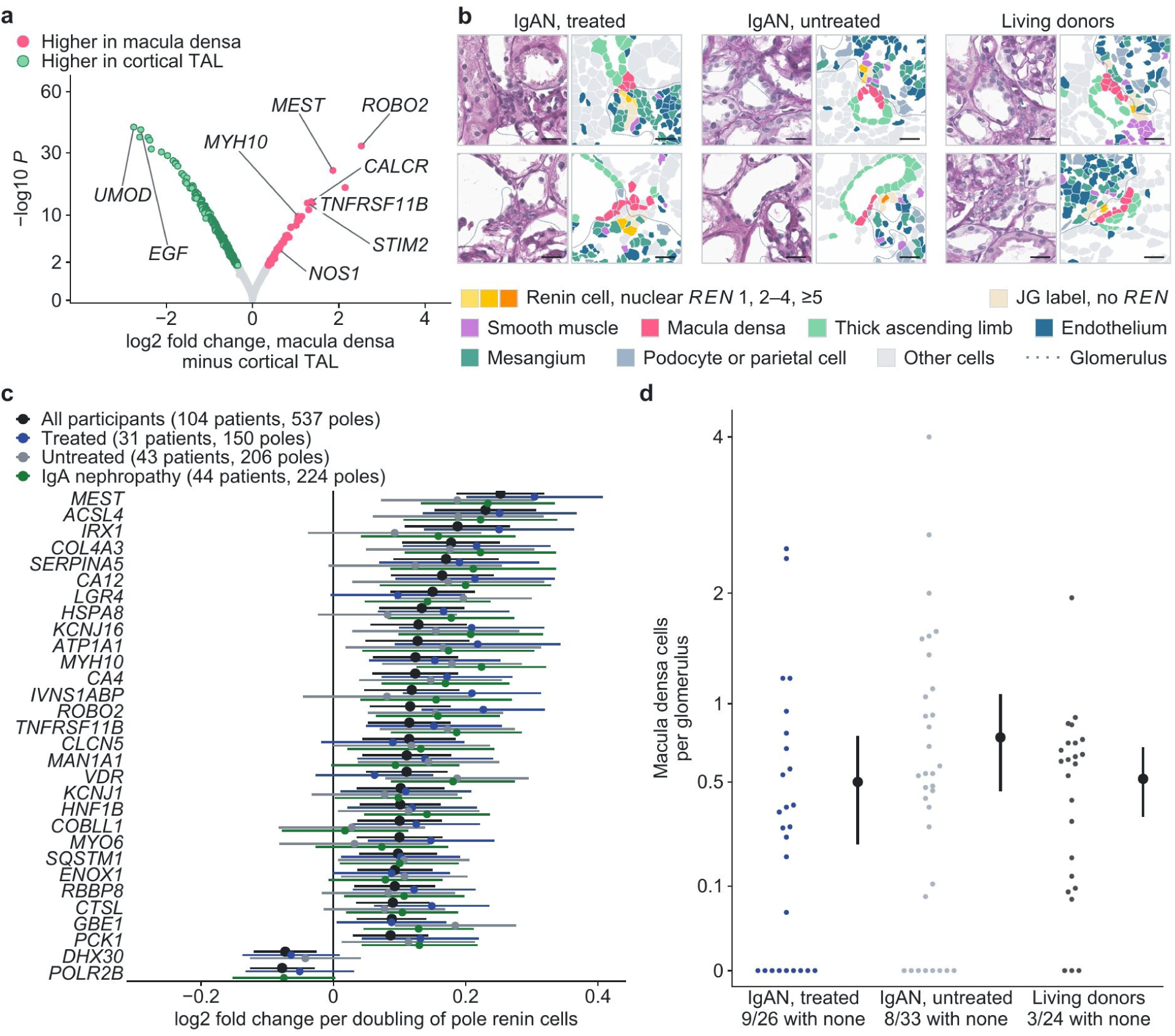
Macula densa and renin cells at the same vascular pole. **a**, Macula densa against cortical thick ascending limb cells (49 participants; colored, genes at *q* < 0.05). **b**, Two vascular poles each of treated patients, untreated patients and living donors with macula densa and renin cells: PAS (left) and segmented cells (right); bars, 20 µm. **c**, Genes of thick ascending limb cells within 15 µm of the glomerulus per doubling of renin cells at the same pole (the 30 genes at a false discovery rate below 10% in all participants), in all participants, in treated and untreated patients and in IgA nephropathy; points and lines, estimates and 95% confidence intervals. **d**, Macula densa cells per glomerulus by group: participants (points) with mean and 95% confidence interval; below, participants without a macula densa cell. IgAN, IgA nephropathy; JG, juxtaglomerular; TAL, thick ascending limb.

Where a vascular pole held more renin cells, the adjacent thick ascending limb expressed more macula densa marker genes: per doubling of renin cells, more *MEST* (log_2_ fold change 0.25; 0.18 to 0.32), *ACSL4*, *COL4A3*, *VDR* and *ROBO2* (30 genes at a false discovery rate below 10%; 537 poles of 104 participants; Figure 4c; Supplementary Table S9), with a positive slope for 11 of the 12 tested genes at least twofold higher in the macula densa, whereas *UMOD* and *EGF* did not fall. The number of macula densa cells was not part of this relation: within a biopsy, poles with more macula densa cells did not hold more renin cells in IgA nephropathy (−0.04; −1.39 to 1.31). The relation was present in treated and untreated patients (*MEST* 0.30 and 0.19, not significantly different) and in IgA nephropathy alone (*MEST* 0.23; 0.13 to 0.34).

Treated patients did not have more macula densa cells per glomerulus than untreated patients (0.50 and 0.76; difference −0.26; −0.69 to 0.16; Figure 4d) or more glomeruli with a macula densa plaque (−6.5 percentage points; −15.3 to 2.2; Supplementary Table S1; Supplementary Figure S6). No gene of macula densa cells reached a false discovery rate of 10%, and no macula densa–renin cell ligand–receptor pair differed by treatment. In thick ascending limb cells next to glomeruli, 41 of the 53 genes that are low in the macula densa were lower in treated patients (37 in the adjusted model), among them *EGF*, *SLC5A3* and *UMOD*, with no single gene at a false discovery rate of 10% (Supplementary Table S8). More renin cells in treated patients were thus not accompanied by more macula densa cells or a difference in any single macula densa gene.

### The same distribution of renin cells along the arteriole

Renin cells of treated patients were more numerous but not farther from the glomerulus. The excess over untreated patients was of similar relative size at every distance from the glomerulus (rate ratio 2.2 at 0–20 µm outside the glomerulus and 1.6 to 2.3 beyond; *P* 0.77 for a difference between distances; Supplementary Figure S7; Supplementary Table S3). Arterioles away from glomeruli with three or more renin cells in the wall were found without RAS blockade as well, in 8 of 31 untreated patients and 5 of 23 living donors against 8 of 24 treated patients, without a difference in density by treatment (Figure 5a,b; Supplementary Figure S8; Supplementary Table S1). Renin cells of such arterioles resembled smooth muscle more than renin cells at the pole of the same patients did, with more *ACTA2* and *LBH* (Figure 5c) and a lower score of the renin-cell program without *REN* (−0.45; −0.78 to −0.12 in IgA nephropathy; Figure 5d).

**Figure 5.**
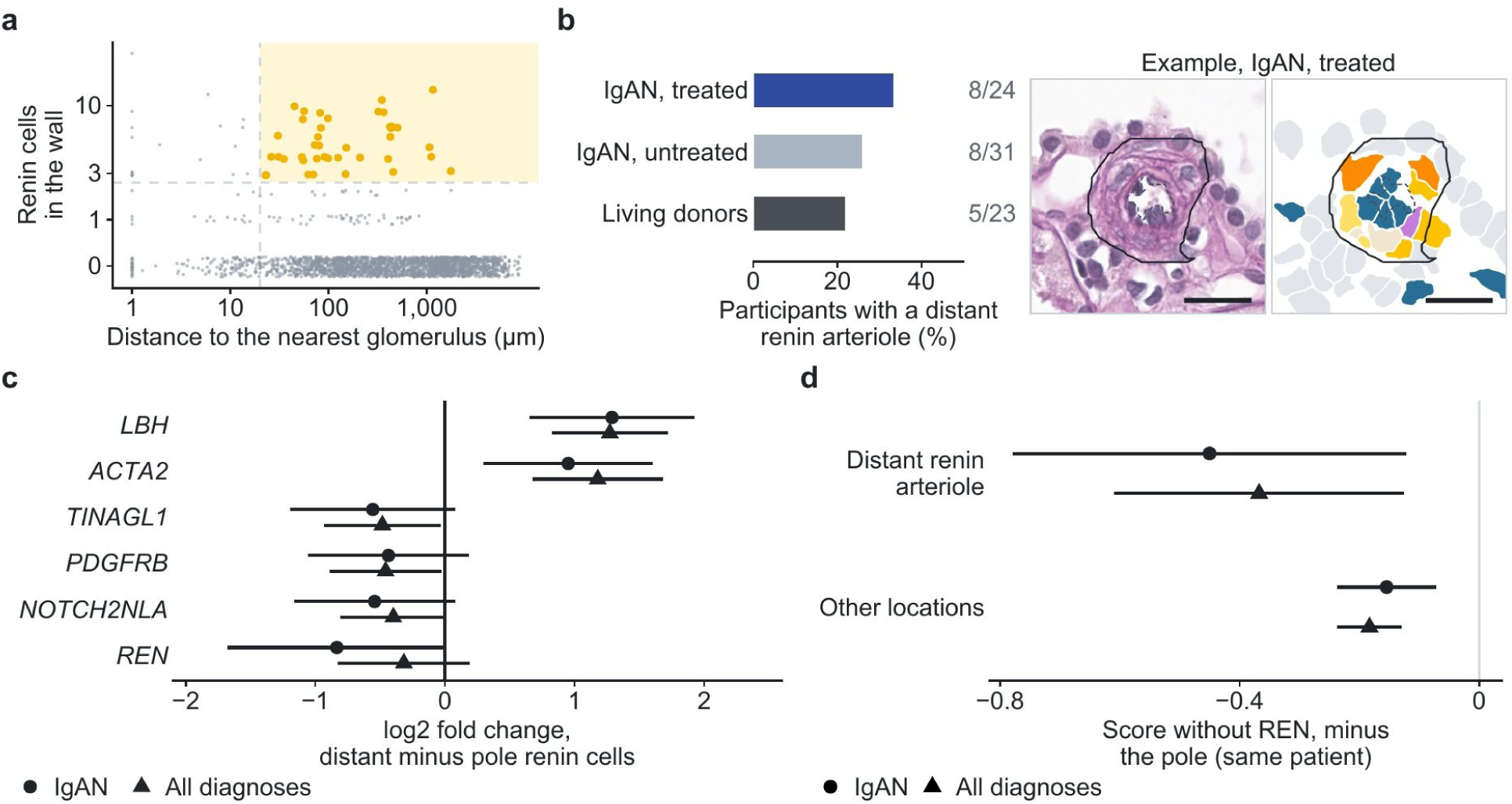
Renin cells along the arteriole. **a**, Renin cells in the wall of every arteriole of the 144 participants with a PAS-registered section against distance to the nearest glomerulus; shaded, distant renin arterioles (more than 20 µm from every glomerulus, three or more renin cells). **b**, Patients with a distant renin arteriole by group (patients at right), and an example from a treated patient: PAS (left) and segmented cells (right); bars, 20 µm. **c**, Renin cells of distant renin arterioles against renin cells at the vascular pole of the same patients: the genes tested in both IgA nephropathy and all diagnoses with *P* < 0.1 in either (every gene tested in Supplementary Data 1). **d**, The same comparison, and renin cells elsewhere in the tissue against renin cells at the pole, on the cell-level score without *REN*. Points and lines (**c**, **d**), estimates and 95% confidence intervals. IgAN, IgA nephropathy.

### Plasma renin, the clinical course and kidney outcomes

In the plasma cohort, 149 patients with IgA nephropathy (74 treated) were followed for a median of 7.9 years, and 49 reached the composite outcome. Plasma renin was not associated with the composite outcome (hazard ratio per standard deviation 0.85; 0.60 to 1.20) or with the eGFR slope, in treated or untreated patients (Supplementary Figure S9).

In the spatial cohort, the renin-cell measures were not associated with eGFR, proteinuria, mean arterial pressure or the Oxford classification at biopsy, or with the composite outcome or the eGFR slope over a median of 8.4 years, with 13 events among the 58 patients followed (Supplementary Table S10). Plasma renin, which rose with renin cells (Figure 2h), was higher in patients with a lower mean arterial pressure at biopsy (−3.97 mmHg per standard deviation; −7.31 to −0.63). Patients with more macula densa cells per glomerulus had lower proteinuria at biopsy, in IgA nephropathy and in non-diabetic glomerular disease (Supplementary Table S10; Supplementary Table S11).

## Discussion

The juxtaglomerular apparatus of patients taking RAS blockade had been described from renin immunostaining and arteriolar morphology in a small series [8] and in single patients [9], and, to our knowledge, not at the level of single cells. In patients with IgA nephropathy, angiotensin receptor blockade was associated with a larger population of juxtaglomerular renin cells, formed by juxtaglomerular cells that transcribed renin, while pole smooth muscle cells did not differ. Renin cells of treated patients had the same renin-cell identity as those of untreated patients, in contrast to the findings in mice. Renin cells increased with macula densa marker genes of the adjacent tubule at each vascular pole and did not lie farther from the glomerulus in treated patients, and plasma renin rose with renin cells.

The difference is in the direction expected from removal of angiotensin II feedback [30] and was of the order of the twofold renin-positive area reported after long-term RAS inhibition in patients [8]. The human data agree with the mouse data only in part. As in mice, renin cells of treated patients were more numerous, and *BGN*, a secreted matrix gene confirmed in the hypertrophied arterioles of mice [8], was higher in renin cells here. Unlike in mice, the renin-cell program of each cell did not differ, the profiles after captopril or renin deletion were not reproduced, and renin cells did not lie farther from the glomerulus. Arterioles away from glomeruli with three or more renin cells were present in every group, living donors included, and their renin cells had a lower score of the renin-cell program without *REN* than renin cells at the pole, consistent with arteriolar smooth muscle cells that express renin [5]. Mice lacking renin are without angiotensin II for life, whereas patients take an angiotensin receptor blocker at clinical doses, so renin cells of patients may respond by number more than by a change of program. The arteriolar hypertrophy reported in patients was seen after more than five years of RAS inhibition [8].

The macula densa signals to renin cells through nitric oxide and prostaglandin E2 [31], and its cyclooxygenase-2 rises with angiotensin-converting enzyme inhibition in rodents [32]. At each pole, renin cells increased with marker genes of the macula densa in the adjacent tubule but not with the number of macula densa cells, in treated and untreated patients alike. The macula densa of treated patients did not differ in cell number or in any single gene, which is consistent with removal of the direct suppression of renin cells by angiotensin II [30].

Concentric arteriolar hypertrophy, which in mice follows the matrix program and the arteriolar accumulation of renin cells, has been proposed as a kidney risk of long-term RAS blockade [11]. Patients taking an angiotensin receptor blocker had more renin cells but, apart from higher secreted matrix genes, none of the changes that precede this hypertrophy in mice. Treated patients had a lower eGFR but more renin cells, a difference that remained with eGFR in the model, and the number of renin cells was not related to kidney function, proteinuria or the kidney course. Plasma renin rose with renin cells with and without treatment, and not with macula densa or pole smooth muscle cells, and may serve as a blood measure of tissue renin cells.

This study has several limitations. This was a retrospective observational study of archival biopsies, in which treatment was not assigned and the indication, dose and duration of RAS blockade could not be controlled, so the differences by treatment cannot be read as effects of the drug. The covariates were measured on treatment, and no tissue from before treatment was available. Renin cells were defined from the transcripts of a targeted panel in one section of each biopsy, without confirmation at the protein level. Kidney outcomes were few among the patients with tissue, which limits the assessment of the kidney course. The participants were Korean patients, mostly from one center, and the findings may not apply to other populations.

## Conclusion

In human IgA nephropathy, RAS blockade is accompanied by a larger population of juxtaglomerular renin cells, in which more juxtaglomerular cells transcribe renin, with the same renin-cell identity and distribution as in untreated patients, in contrast to the findings in mice. At each vascular pole, renin cells increase with macula densa marker genes of the adjacent tubule, and plasma renin rises with tissue renin cells.

## Acknowledgments

The biospecimens and data used for this study were provided by the Biobank of Seoul National University Hospital, a member of the Korea Biobank Network, and by the Korea National Institute of Health (KNIH) research project (project No. KBN5_A03).

## Data Availability

Gene-, cell-type- and model-level summary data underlying the figures and tables are provided in the Supplementary Tables and Supplementary Data 1. Individual-level spatial transcriptomic and clinical data are not publicly available because the archival biopsy tissue was collected under consent for research in Korea, and neither the consent nor the approval of the Institutional Review Board of Seoul National University Hospital covers transfer of individual-level data outside Korea. De-identified data can be shared through a collaboration agreement with the KORNERSTONE investigators, subject to approval by the Institutional Review Board and the Biobank of Seoul National University Hospital; requests should be addressed to the corresponding authors. KPMP single-nucleus data are available from the KPMP data portal (https://atlas.kpmp.org), and the mouse data sets are in the Gene Expression Omnibus (GSE180873, GSE42712, GSE17138, GSE146386, GSE157699).

## Code Availability

The analysis code is deposited at Zenodo (https://doi.org/10.5281/zenodo.24308485) and is available on request.

## Author Contributions

S.G.H. and S.P. conceived the study. S.G.H. performed the spatial, external-cohort and statistical analyses and drafted the manuscript. A.K. curated clinical data, interpreted the results, finalized the manuscript and coordinated its submission and revision. M.K. developed the kidney structure segmentation model and produced the PAS outlines. D.K. performed cell-type annotation of the spatial transcriptomic data. J.H.J. and J.S.L. contributed to data generation and reviewed the final manuscript. H.K., J.-I.O., J.L., M.W.K., Y.S., H.L., S.S.H., Y.K., J.K., J.H.L., J.P.L., J.I.P., J.T.P., J.N.A., K.-C.M. and H.J.K. contributed to participant enrollment and data generation and reviewed the final manuscript. Y.-G.K. supervised image analysis and segmentation. S.P. curated clinical data, drafted the manuscript and supervised the analyses. D.K.K. established the cohort and supervised the study. All authors revised the manuscript and approved the final version.

