## Supplemental information for "Spatial transcriptomics of juxtaglomerular renin cells under angiotensin receptor blockade in human IgA nephropathy"

#### Table of Contents

**Supplementary Methods** 3
Cohort and clinical data 3
Spatial transcriptomics, glomeruli and renin cells 3
Renin cells by distance from the glomerulus 4
Aggregate vascular pole 4
Arterioles 4
Statistical models 4
Macula densa and cells of the vascular pole 5
Expression analyses 5
Tissue panels 6
External data and plasma 6
Kidney course in the plasma cohort 6
Clinical course in the spatial cohort 6
Mouse renin-cell gene sets and profiles 7
Software 7

**Supplementary Tables**
Supplementary Table S1. Renin cells and the vascular pole by RAS blockade in IgA nephropathy 8
Supplementary Table S2. Renin-cell definitions 9
Supplementary Table S3. Location of renin cells and renin cells by distance to the nearest glomerulus 10
Supplementary Table S4. Sensitivity analyses, the Kidney Precision Medicine Project and plasma renin 12
Supplementary Table S5. Renin cells against arteriolar smooth muscle 14
Supplementary Table S6. Renin cells of treated against untreated patients 15
Supplementary Table S7. Mouse renin-cell gene sets and profiles in renin cells of treated patients 17
Supplementary Table S8. Macula densa markers, macula densa expression by treatment and ligand–receptor pairs 18
Supplementary Table S9. Thick ascending limb genes at the vascular pole and renin cells at the same pole 20
Supplementary Table S10. Juxtaglomerular measures, the clinical state at biopsy and the kidney course in IgA nephropathy 21
Supplementary Table S11. Juxtaglomerular measures, the clinical state at biopsy and the kidney course in non-diabetic glomerular disease 22

**Supplementary Figures**
Supplementary Figure S1. Cells of the vascular pole by treatment 23
Supplementary Figure S2. Renin-cell calls and their quality 24
Supplementary Figure S3. Renin nuclei in the Kidney Precision Medicine Project atlas 26
Supplementary Figure S4. Mouse renin-cell gene sets and profiles 27
Supplementary Figure S5. Vascular poles by group 28
Supplementary Figure S6. Macula densa plaques 29
Supplementary Figure S7. Renin cells by distance from the glomerulus 30
Supplementary Figure S8. Distant renin arterioles 31
Supplementary Figure S9. Plasma renin and kidney outcomes in the plasma cohort 32

**Supplementary References** 33

**Supplementary Data 1 (separate Excel file)**
Gene-level and model-level results behind the figures and tables; the README sheet lists every sheet and the display it supports.

### Supplementary Methods

#### Cohort and clinical data

The spatial cohort comprised the 154 participants with a primary single-cell-level spatial transcriptomics section in the KORNERSTONE biobank network: sequencing batch 1 held 96 sections (IgA nephropathy 24, minimal change disease 14, membranous nephropathy 10, diabetic kidney disease 24 and all 24 living donors) and batch 2 held 58. In IgA nephropathy, 7 of 24 patients of batch 1 and 19 of 36 of batch 2 were treated (Table 1). Every participant of the spatial cohort had the biopsy at Seoul National University Hospital; of the 225 patients of the plasma cohort, 198 had it there and 27 at other KORNERSTONE hospitals. Plasma collected at biopsy was the sample of the plasma registry drawn within 7 days of the biopsy, the nearest one when there were several; it was drawn on the day of the biopsy for 204 of the 225 patients of the plasma cohort and within 3 days for the others, and 98 participants of the spatial cohort, 51 of them with IgA nephropathy, had such a sample. Medication before biopsy came from the clinical research record of the cohort. Mean arterial pressure was the diastolic pressure plus one third of the pulse pressure at biopsy, and eGFR was computed with the 2021 CKD-EPI equation from the serum creatinine at biopsy (living donors, before donation). In IgA nephropathy, eGFR at biopsy was computed from the serum creatinine of the hospital laboratory record nearest the biopsy, with age counted from the first day of the birth month.

#### Spatial transcriptomics, glomeruli and renin cells

Sections of formalin-fixed, paraffin-embedded biopsies were profiled with the Xenium Prime 5K panel (5,101 genes), cells were segmented and annotated with the cohort atlas labels, and the PAS-stained section was registered to the cell coordinates by landmark alignment with local correction. Transcripts with a quality value of at least 20 were counted in the nucleus of each cell. Glomeruli were density-based clusters (DBSCAN [S1]; radius 40 µm, at least 10 cells) of podocytes, mesangial cells and parietal epithelial cells, with the convex hull of each cluster as its boundary; the distance of a cell from the glomerulus was its distance from the nearest hull of the section, zero inside a hull. Cell identities, the juxtaglomerular label among them, came from the cohort annotation of the spatial data, which defined 24 kidney populations by graph clustering of a 50-dimensional latent expression representation of all sections and review of marker genes. The juxtaglomerular label marked cells with high *REN*, *ABCC8*, *KCNJ2*, *WFDC1* and *KCNA5*. Nuclear transcripts of *REN* (56.3% of cells with the juxtaglomerular label against 0.7% of cells with the vascular smooth muscle label), *TRPM3* (13.4% against 0.9%), *KCNJ2* and *MECOM* were detected more often in labeled cells, and those of *ACTA2* (18.3% against 38.5%) and *VIM* less often. The label did not require a nuclear *REN* transcript, and 6,305 of the 14,444 labeled cells (43.7%) carried none (Supplementary Figure S2a,b).

A renin cell had the juxtaglomerular label and at least one nuclear *REN* transcript; at least three transcripts, with or without the label, and the label with any *REN* count were the other definitions (Supplementary Table S2; Supplementary Figure S2c). Each renin cell was assigned to its nearest glomerulus and counted at that glomerulus when it lay within 20 µm of it; renin cells per glomerulus were these counts summed over the glomeruli of a participant and divided by their number. Mural cells of the vascular pole were the renin cells, smooth muscle cells and labeled cells without a nuclear *REN* transcript within 20 µm of a glomerulus, and the renin share of pole mural cells was the renin cells among them. To examine transcript spillover, the share of cells of other lineages (podocytes, parietal epithelial, mesangial, endothelial and thick ascending limb cells) within 20 µm of a glomerulus that carried a nuclear *REN* transcript was compared by treatment, together with the renin cells per glomerulus that spillover at the same rate would add. The nuclear transcripts per mural-lineage cell within 20 µm, the labeled cells per glomerulus and the share of labeled cells with a nuclear *REN* transcript are given by sequencing batch and group (Supplementary Table S2; Supplementary Figure S2d,e).

#### Renin cells by distance from the glomerulus

The renin-cell density of a distance band was the number of renin cells in the band over the tissue area of the band, with tissue area taken from a 4-µm grid around the cell centroids of the section. The renin-cell counts of the bands inside the glomerular outline, 0–20, 20–50, 50–100, 100–300 and beyond 300 µm were modeled together by negative binomial generalized estimating equations with the log tissue area of the band as offset, a treatment term for each band, age, sex and batch, patient as the cluster and bias-reduced sandwich errors [S2]; one test asked whether the rate ratio differed between bands (Supplementary Table S3; Supplementary Figure S7).

#### Aggregate vascular pole

For each glomerulus with at least three mural-lineage cells within 20 µm, the pole axis ran from the centroid of the glomerulus to the centroid of the mural cells within 20 µm that were not renin cells. The renin cells of each glomerulus were placed relative to the centroid of the glomerulus, scaled by its equivalent radius and rotated so that the axis pointed up, and the map of a group gave these cells per glomerulus and per unit of the squared radius after Gaussian smoothing (Figure 2a); glomeruli whose mural cells within 20 µm were all renin cells had no axis.

#### Arterioles

Arterioles were outlined on the PAS images by an instance segmentation model (Mask R-CNN [S3] with convolutional block attention modules [S4]; mask average precision 0.839 on held-out patches) and identified among the vascular profiles from their PAS morphology and the identity of the cells in the wall. Arterioles with an outer diameter of up to 100 µm were analyzed. A distant renin arteriole lay more than 20 µm from every glomerulus of its section and held at least three renin cells in its wall (Supplementary Figure S8).

#### Statistical models

Participant measures were regressed on RAS blockade with age, sex and batch (basic model) or also mean arterial pressure, log UPCR and eGFR (adjusted model), with HC3 heteroskedasticity-consistent errors [S5]; in non-diabetic glomerular disease the models added a term for diagnosis. The 39 patients without hypertension, the 48 patients without a calcium-channel blocker and each sequencing batch (without the batch term) were also analyzed alone (Supplementary Table S4). Renin-cell counts were also modeled by negative binomial regression with the log number of glomeruli as offset and sandwich errors, the exponentiated treatment term being the rate ratio. Within a biopsy, the poles holding at least one macula densa cell were split at the participant's median number of macula densa cells, and renin cells within 20 µm were compared between the upper and the lower half with participant fixed effects, among participants with poles in both halves.

#### Macula densa and cells of the vascular pole

Thick ascending limb cells within 15 µm of a glomerulus and within 40 µm of its vascular pole formed the macula densa. The vascular pole of a glomerulus was the centroid of its smooth muscle cells and juxtaglomerular-labeled cells without a nuclear *REN* transcript within 20 µm, the same point that defined the pole axis of the aggregate map (Figure 2a); glomeruli with fewer than three such cells had no pole. Macula densa cells of one glomerulus were grouped into plaques by single-linkage clustering at 10 µm (Supplementary Figure S6). Cortical thick ascending limb cells were those more than 50 and up to 300 µm from a glomerulus. Pole smooth muscle comprised mural-lineage cells without the juxtaglomerular label within 20 µm of a glomerulus. Extraglomerular mesangial cells were mesangial cells within 30 µm of the center of a macula densa plaque. Cells of every class within 20 µm of a glomerulus and 40 µm of its pole were counted per pole and compared by treatment as participant means (Supplementary Figure S1).

#### Expression analyses

Nuclear transcripts of each cell population of a participant were summed into a pseudobulk library and analyzed with limma-voom, with the mean transcripts per cell of the library as a covariate; populations with fewer than 10 renin cells or 20 other cells were left out, and genes were kept by edgeR filterByExpr with the compared groups. Renin cells were compared with the arteriolar smooth muscle of the same patients with participant blocks, and between treated and untreated patients with the covariates of the basic or adjusted model (Supplementary Tables S5 and S6). Renin cells of distant renin arterioles were compared with the renin cells within 20 µm of a glomerulus of the same patients, with participant blocks (Figure 5c); the pseudobulk comparison of renin cells by location required at least three renin cells of a participant in distant renin arterioles and five at the pole, and the macula densa comparisons at least 10 macula densa cells. Marker genes of the macula densa were the genes at $q < 0.05$ with at least a twofold difference between the macula densa and cortical thick ascending limb, and their direction was counted in the slopes per vascular pole and in the comparison by treatment. Gene sets (MSigDB Hallmark, Gene Ontology Biological Process and Reactome; 10 to 300 genes among the tested genes) were tested with cameraPR on the moderated *t* statistics of each comparison (Supplementary Data 1).

At each vascular pole with at least three thick ascending limb cells within 15 µm of the glomerulus, these cells were summed into one library, participants with at least two such poles were kept, and each library was related to $\log_2$(1 + renin cells within 20 µm of the same glomerulus) with participant blocks and transcript yield as a covariate (537 poles of 104 participants; Supplementary Table S9). The same model was fitted in treated and in untreated patients and, with the interaction of the renin-cell term and RAS blockade, in all patients with a medication record (Figure 4c). Ligand–receptor pairs from CellChatDB v2 [S6] with the ligand detected in macula densa cells and the receptor in pole renin cells were scored at each pole as the mean ligand expression of the macula densa cells times the mean receptor expression of the renin cells, each on $\log_2$(1 + 100 × nuclear count / nuclear transcripts of the cell), and the score was compared by treatment (Supplementary Table S8).

The cell-level renin score projected each cell's log-normalized nuclear counts onto the $\log_2$ fold changes of the genes at $q < 0.05$ between renin cells and arteriolar smooth muscle, scaled so that the average smooth muscle cell scored 0 and the average renin cell 1; a version without *REN* was used to compare cells by location. Participant medians of the score were compared by treatment in the basic and adjusted models, and locations within participants with participant fixed effects.

#### Tissue panels

Tissue panels show the cell outlines of the original Xenium output filled by the identities of the analysis, with renin cells shaded by nuclear *REN* count (1, 2–4, 5 or more transcripts), beside the registered PAS image of the same window. Supplementary Figure S8 shows every distant renin arteriole of patients with IgA nephropathy and of living donors.

#### External data and plasma

KPMP single-nucleus counts and the open-access field for RAS blockade were used for 116 participants [S7]; renin nuclei were those of subclass REN, and vascular mural nuclei those of REN, VSMC, VSMC/P and dVSMC (level-2 annotation). Renin nuclei per 1,000 nuclei and per vascular mural nuclei were compared by RAS blockade with least squares, without and with adjustment for age-bin midpoint, sex and enrollment category, in all participants and in chronic kidney disease, and renin-nucleus pseudobulk libraries were compared by RAS blockade with edgeR [S8] (Supplementary Figure S3). For the 50 renin-cell genes with the smallest *P* across diagnoses here, the number measured in KPMP that had the same sign was compared with 50% by a binomial test (Supplementary Table S4). For renin-cell identity, the KPMP renin and vascular smooth muscle libraries of the 121 participants with both were summed per cell type, and their $\log_2$ counts per million were compared with the differences of renin cells against smooth muscle here (Spearman correlation). Plasma renin was compared between treated and untreated patients with age and sex as covariates, and tissue measures were regressed on plasma renin (per NPX) with age, sex, sequencing batch and RAS blockade (Supplementary Table S4).

#### Kidney course in the plasma cohort

eGFR values were computed with the 2021 CKD-EPI equation from serum creatinine, one value per day. The SNUH laboratory record of the plasma cohort holds reported eGFR values only, so each value was converted back to creatinine with the exact inverse of its equation at the age on the day, and the serum creatinine at biopsy of the plasma registry was added on its day; for the 51 patients also in the spatial cohort, the measured creatinine values of the spatial cohort were used. The eGFR at biopsy was the value nearest the biopsy. The composite outcome was the first eGFR after the biopsy at or below 60% of the eGFR at biopsy or below 15 mL/min/1.73 $m^2$ that was confirmed by a value at or below 60% at least 28 days later, unless values were recorded within those 28 days and all had returned to 80% or more; a decline without a value 28 days or more later counted when it was the last value or when not all later values had returned to 80% or more. Kidney failure was the second eGFR below 15 mL/min/1.73 $m^2$, values after it were not used, and the composite took whichever came first. Patients with at least two eGFR values after the biopsy, the last at least 3 months after it were followed and censored at their last value. The eGFR slope was the least-squares slope of all values from the biopsy to kidney failure (at least three over at least one year). The composite outcome was analyzed by Cox models with plasma renin per standard deviation, age, sex and RAS blockade (basic) and also eGFR and log UPCR at biopsy (adjusted), in all patients and within each treatment group, and with the first eGFR at or below 60% without confirmation as the outcome. Kaplan–Meier curves compared plasma renin above and below the median within each treatment group (log-rank test; Supplementary Figure S9).

#### Clinical course in the spatial cohort

Patients of the spatial cohort with non-diabetic glomerular disease (IgA nephropathy, minimal change disease, membranous nephropathy and focal segmental glomerulosclerosis) had the measured serum creatinine values of the hospital laboratory record, from which daily eGFR values were computed with the 2021 CKD-EPI equation and age counted from the first day of the birth month; the composite outcome, kidney failure and follow-up were defined as in the plasma cohort. The eGFR slope was the measure-by-year term of a linear mixed model of every eGFR value from biopsy to kidney failure, with a random intercept and slope per patient and a year term for each covariate. The UPCR at 6 months was the random-urine value nearest day 183 within days 92 to 273, expressed as the $\log_2$ ratio to the UPCR at biopsy; partial remission was any value within 12 months at or below half the UPCR at biopsy and below 1 g/g. Each juxtaglomerular measure and plasma renin entered per standard deviation of the cohort, with age, sex, sequencing batch, RAS blockade, eGFR and log UPCR at biopsy as covariates except the one that was the outcome, in least squares models with heteroskedasticity-consistent errors, Cox models and logistic regression; in non-diabetic glomerular disease the models added a diagnosis term and the Cox models were stratified by diagnosis (Supplementary Table S10; Supplementary Table S11). In IgA nephropathy, each measure was also related to the Oxford classification by logistic (M, E, S) and proportional odds (T, C) models adjusted for age, sex, sequencing batch and RAS blockade (Supplementary Data 1).

#### Mouse renin-cell gene sets and profiles

Gene sets were defined in the source data and mapped to one-to-one human orthologs (Ensembl release 116 [S9]); *Ren1* and *REN* were left out. The matrix-secretory gene set held the genes higher in renin-lineage cells after *Ren1c* deletion than in wild-type renin cells in the published single-cell analysis [S10] (446 genes, 133 on the panel), with its secreted-protein subset (58, 22 on the panel); the endocrine identity set held the genes higher in wild-type renin cells (134, 32 on the panel). Captopril: renin cells of adult mice given captopril with a low-salt diet for 2 weeks against untreated adult renin cells (GSE42712, GSE17138 [S11]). Recruitment: renin cells recruited by captopril with a low-salt diet for 1 week (GSE146386 [S12]) against renin cells of sham-operated mice (GSE157699); low perfusion pressure: renin cells of the kidney downstream of an aortic coarctation against the contralateral kidney (GSE157699 [S13]). Gene sets of the captopril, recruitment and low-pressure data were the genes at unadjusted $P < 0.01$ with an absolute $\log_2$ fold change of at least 1. For each set, the mean moderated *t* of its measured genes minus the mean of the other genes, in renin cells of treated against untreated patients, was compared with 10,000 random sets of the same size, and each set was also tested with limma camera [S14]. Whole profiles were compared by the Spearman correlation of the human and mouse statistics over the genes measured in both species, and the gene sets were also tested in the comparison of renin cells with arteriolar smooth muscle (Supplementary Table S7; Supplementary Figure S4).

#### Software

Analyses used Python 3.14 (pandas, NumPy, SciPy, statsmodels and shapely), R 4.6 (limma, edgeR, ggplot2 and patchwork) and DuckDB for tables.

#### Supplementary Tables

##### Supplementary Table S1. Renin cells and the vascular pole by RAS blockade in IgA nephropathy

*A. Renin cells*

| | IgA nephropathy (59 patients, 26 treated) | | | | |
|---|---|---|---|---|---|
| Measure | RAS blockade, basic | *P* | *q* | RAS blockade, adjusted | Non-diabetic glomerular disease (89; 40 treated) |
| Renin cells per glomerulus | 1.30 (0.42 to 2.18) | 0.004 | 0.013 | 1.34 (0.28 to 2.40) | 1.37 (0.64 to 2.09) |
| Renin cells per glomerulus, rate ratio | 2.40 (1.55 to 3.70) | <0.001 | – | 2.39 (1.37 to 4.13) | 2.44 (1.70 to 3.48) |
| Renin cells per $mm^2$ of tissue | 6.8 (0.3 to 13.2) | 0.040 | 0.089 | 5.0 (−0.8 to 10.8) | 8.1 (2.9 to 13.4) |
| Renin share of pole mural cells, % | 12.4 (5.3 to 19.5) | <0.001 | 0.008 | 12.7 (4.5 to 20.9) | 11.7 (5.9 to 17.6) |
| Labeled pole cells with nuclear *REN*, % | 19 (7 to 31) | 0.002 | 0.009 | 19 (6 to 32) | 20 (11 to 30) |
| Distant renin arterioles per $mm^2$ | 0.03 (−0.08 to 0.14) | 0.596 | 0.670 | 0.01 (−0.09 to 0.11) | 0.02 (−0.07 to 0.10) |

*B. Macula densa and vascular pole*

| | IgA nephropathy (59 patients, 26 treated) | | | | |
|---|---|---|---|---|---|
| Measure | RAS blockade, basic | *P* | *q* | RAS blockade, adjusted | Non-diabetic glomerular disease (89; 40 treated) |
| Macula densa cells per glomerulus | −0.26 (−0.69 to 0.16) | 0.211 | 0.316 | −0.17 (−0.61 to 0.27) | −0.15 (−0.46 to 0.16) |
| Glomeruli with a macula densa plaque, % | −6.5 (−15.3 to 2.2) | 0.138 | 0.248 | −4.5 (−13.6 to 4.5) | −4.3 (−11.0 to 2.4) |
| Pole smooth muscle cells per glomerulus | 0.02 (−1.61 to 1.65) | 0.981 | 0.981 | −0.26 (−1.99 to 1.47) | 0.04 (−1.17 to 1.25) |
| Extraglomerular mesangial cells per glomerulus | −0.18 (−0.72 to 0.35) | 0.491 | 0.631 | −0.10 (−0.66 to 0.46) | −0.20 (−0.58 to 0.17) |

Treated minus untreated (95% CI); rate ratio of renin cells per glomerulus from a negative binomial model with the number of glomeruli as exposure (not in the *q* family). Basic, age, sex and sequencing batch; adjusted, also mean arterial pressure, log UPCR and eGFR; non-diabetic glomerular disease, IgA nephropathy, minimal change disease, membranous nephropathy and focal segmental glomerulosclerosis (patients; treated) with a diagnosis term. Labeled pole cells, juxtaglomerular-labeled cells within 20 µm of a glomerulus; macula densa, thick ascending limb cells within 15 µm of a glomerulus and 40 µm of its vascular pole; glomeruli with a plaque, as a percentage of the glomeruli of the section. Patients differ from the heading in three rows: renin cells per $mm^2$ of tissue, IgA nephropathy 60 (26 treated); labeled pole cells, 56 (24 treated); distant renin arterioles, patients with a section registered to PAS histology (IgA nephropathy 55, 24 treated; non-diabetic glomerular disease 82, 36 treated). *q*, Benjamini–Hochberg over the 9 rows other than the rate ratio; –, not estimated. eGFR, estimated glomerular filtration rate (CKD-EPI 2021); PAS, periodic acid–Schiff; RAS, renin–angiotensin system; UPCR, urine protein-to-creatinine ratio.

#### Supplementary Table S2. Renin-cell definitions

*A. Juxtaglomerular-labeled cells by nuclear REN transcripts*

| Group | Cells, *n* | % |
|---|---|---|
| 0 transcripts | 6,305 | 43.7 |
| 1 transcript | 1,654 | 11.5 |
| 2 transcripts | 1,129 | 7.8 |
| ≥3 transcripts | 5,356 | 37.1 |
| Mural cells without the label with ≥3 transcripts | 244 | – |

*B. Labeled cells without nuclear REN by distance to the nearest glomerulus*

| Distance | Without *REN*, *n* | % |
|---|---|---|
| 0–20 μm | 2,088 of 5,525 | 38 |
| 20–50 μm | 565 of 1,935 | 29 |
| 50–100 μm | 526 of 1,248 | 42 |
| >100 μm | 3,111 of 5,719 | 54 |
| No glomerulus in section | 15 of 17 | 88 |

*C. Renin cells per glomerulus, IgA nephropathy, treated minus untreated (basic model)*

| Definition | Difference (95% CI) |
|---|---|
| Label and ≥1 transcript (primary) | 1.30 (0.42 to 2.18) |
| Mural-lineage cell with ≥3 transcripts, label or not (secondary) | 0.92 (0.31 to 1.54) |
| Juxtaglomerular label, any *REN* count | 1.26 (0.10 to 2.42) |

*D. Calls by sequencing batch, median (lower, upper quartile)*

| Batch and group | Participants | Transcripts per mural cell | Labeled cells per glomerulus | Labeled cells with *REN*, % |
|---|---|---|---|---|
| Batch 1, IgA nephropathy, treated | 7 | 26 (20, 68) | 2.90 (2.28, 3.67) | 67 (60, 75) |
| Batch 1, IgA nephropathy, untreated | 17 | 30 (15, 40) | 2.00 (0.59, 3.08) | 50 (40, 64) |
| Batch 1, Living donors | 24 | 38 (30, 60) | 1.04 (0.43, 1.86) | 57 (50, 69) |
| Batch 2, IgA nephropathy, treated | 19 | 27 (21, 38) | 3.40 (1.18, 4.63) | 68 (58, 74) |
| Batch 2, IgA nephropathy, untreated | 16 | 34 (15, 46) | 2.03 (1.19, 2.39) | 44 (21, 57) |

Nuclear transcripts with a quality value of at least 20. Of the 261,766 mural-lineage cells of the 154 sections, 14,444 carried the juxtaglomerular label. Labeled cells without a nuclear *REN* transcript were assigned to neither renin cells nor smooth muscle. C, the secondary definition does not require the label. D, mural-lineage cells within 20 μm of a glomerulus; transcripts per cell, nuclear transcripts of all genes; participants with at least one glomerulus.

**Supplementary Table S3. Location of renin cells and renin cells by distance to the nearest glomerulus**

*A. Location of renin cells*

| Group and place | Renin cells, *n* | % |
|---|---|---|
| All participants (153 participants; 8,139 renin cells) | | |
| Within 20 µm of a glomerulus | 3,437 | 42.2 |
| Arteriolar wall beyond 20 µm | 381 | 4.7 |
| Elsewhere, 20–100 µm from a glomerulus | 1,922 | 23.6 |
| Elsewhere, beyond 100 µm | 2,399 | 29.5 |
| IgA nephropathy, treated (26 participants; 1,693 renin cells) | | |
| Within 20 µm of a glomerulus | 752 | 44.4 |
| Arteriolar wall beyond 20 µm | 130 | 7.7 |
| Elsewhere, 20–100 µm from a glomerulus | 354 | 20.9 |
| Elsewhere, beyond 100 µm | 457 | 27.0 |
| IgA nephropathy, untreated (34 participants; 1,580 renin cells) | | |
| Within 20 µm of a glomerulus | 609 | 38.5 |
| Arteriolar wall beyond 20 µm | 117 | 7.4 |
| Elsewhere, 20–100 µm from a glomerulus | 440 | 27.8 |
| Elsewhere, beyond 100 µm | 414 | 26.2 |
| Living donors (24 participants; 1,410 renin cells) | | |
| Within 20 µm of a glomerulus | 472 | 33.5 |
| Arteriolar wall beyond 20 µm | 48 | 3.4 |
| Elsewhere, 20–100 µm from a glomerulus | 333 | 23.6 |
| Elsewhere, beyond 100 µm | 557 | 39.5 |

*B. Renin cells per mm² of tissue in each distance band from the nearest glomerulus*

| | Median (lower, upper quartile) | | | Rate ratio, a separate model per band | |
|---|---|---|---|---|---|
| Distance | IgAN, treated | IgAN, untreated | Living donors | Basic | Adjusted |
| 0–20 µm | 106.1 (48.7, 160.5) | 39.2 (11.2, 89.1) | 29.6 (14.5, 58.1) | 2.33 (1.54 to 3.51) | 2.23 (1.32 to 3.78) |
| 20–50 µm | 30.8 (1.7, 106.5) | 26.8 (4.0, 51.9) | 30.1 (13.0, 54.4) | 1.42 (0.72 to 2.79) | 1.18 (0.56 to 2.47) |
| 50–100 µm | 9.8 (0.9, 16.0) | 2.5 (0.0, 20.9) | 3.9 (1.3, 15.5) | 2.14 (1.21 to 3.77) | 1.56 (0.83 to 2.90) |
| 100–300 µm | 3.4 (0.0, 6.6) | 1.7 (0.0, 4.7) | 3.8 (1.6, 8.6) | 2.98 (1.37 to 6.46) | 1.83 (0.90 to 3.71) |
| >300 µm | 1.2 (0.1, 5.9) | 0.8 (0.0, 3.0) | 3.1 (1.0, 6.5) | 1.21 (0.49 to 2.94) | 1.22 (0.44 to 3.36) |

*Supplementary Table S3, continued: C. Rate ratio of treated against untreated IgA nephropathy by band, one model over all bands*

| Band | Patients | Rate ratio (95% CI) | *P* |
|---|---|---|---|
| Inside the glomerular outline | 59 | 3.21 (1.54 to 6.66) | 0.002 |
| 0–20 µm outside the outline | 59 | 2.25 (1.42 to 3.56) | <0.001 |
| 20–50 µm | 59 | 1.73 (0.92 to 3.23) | 0.082 |
| 50–100 µm | 59 | 1.85 (0.98 to 3.49) | 0.054 |
| 100–300 µm | 59 | 2.28 (0.95 to 5.46) | 0.064 |
| >300 µm | 59 | 1.61 (0.67 to 3.86) | 0.278 |
| Treatment-by-band terms, joint *F* test | 59 | – | 0.766 |

A, every renin cell of the 153 sections that held one. B, renin cells per $mm^2$ of the tissue in each band; rate ratio, negative binomial regression with band area as exposure, a separate model for each band (models as in Supplementary Table S1). C, one model over all bands with band-by-treatment terms (generalized estimating equations within patient, bias-reduced sandwich errors, *t* and *F* references on 44 degrees of freedom). IgAN, IgA nephropathy.

**Supplementary Table S4. Sensitivity analyses, the Kidney Precision Medicine Project and plasma renin**

*A. Renin cells per glomerulus, IgA nephropathy unless stated, treated minus untreated*

| Analysis | Patients | Difference (95% CI) | *P* |
|---|---|---|---|
| Basic model | 59 | 1.30 (0.42 to 2.18) | 0.004 |
| Adjusted model | 59 | 1.34 (0.28 to 2.40) | 0.014 |
| Patients without hypertension (9 treated, 30 untreated), basic | 39 | 1.66 (−0.93 to 4.24) | 0.201 |
| Patients without a calcium-channel blocker (17 treated, 31 untreated), basic | 48 | 1.25 (0.00 to 2.51) | 0.050 |
| At least three nuclear *REN* transcripts, with or without the label, basic | 59 | 0.92 (0.31 to 1.54) | 0.004 |
| Sequencing batch 1 (age and sex) | 24 | 0.86 (−0.26 to 1.98) | 0.122 |
| Sequencing batch 2 (age and sex) | 35 | 1.61 (0.34 to 2.88) | 0.014 |
| Non-diabetic glomerular disease (89 patients, 40 treated), basic | 89 | 1.37 (0.64 to 2.09) | <0.001 |

*B. Renin cells per glomerulus, IgA nephropathy, treated to untreated*

| Analysis | Patients | Rate ratio (95% CI) | *P* |
|---|---|---|---|
| Glomeruli as exposure, basic | 59 | 2.40 (1.55 to 3.70) | <0.001 |
| Glomeruli as exposure, adjusted | 59 | 2.39 (1.37 to 4.13) | 0.002 |
| Patients without hypertension, basic | 39 | 2.34 (1.24 to 4.38) | 0.008 |
| Patients without a calcium-channel blocker, basic | 48 | 2.16 (1.31 to 3.55) | 0.002 |
| At least three nuclear *REN* transcripts, with or without the label, basic | 59 | 2.82 (1.68 to 4.74) | <0.001 |
| Sequencing batch 1 (age and sex) | 24 | 1.59 (1.03 to 2.45) | 0.035 |
| Sequencing batch 2 (age and sex) | 35 | 2.96 (1.70 to 5.15) | <0.001 |
| Pooled over batches (heterogeneity *P* = 0.081) | 59 | 2.01 (1.43 to 2.83) | <0.001 |

*C. Kidney Precision Medicine Project, on against not on RAS blockade*

| Comparison | Participants | Difference (95% CI) | *P* |
|---|---|---|---|
| All participants, renin nuclei per 1,000 nuclei, unadjusted | 116 | 0.58 (0.11 to 1.06) | 0.016 |
| All participants, renin nuclei per 1,000 nuclei, adjusted | 116 | 0.65 (0.12 to 1.18) | 0.016 |
| All participants, renin nuclei, % of vascular mural nuclei, unadjusted | 115 | 1.66 (−3.26 to 6.58) | 0.506 |
| All participants, renin nuclei, % of vascular mural nuclei, adjusted | 115 | 4.86 (−0.34 to 10.05) | 0.066 |
| Chronic kidney disease, renin nuclei per 1,000 nuclei, unadjusted | 72 | 0.69 (0.03 to 1.34) | 0.040 |
| Chronic kidney disease, renin nuclei per 1,000 nuclei, adjusted | 72 | 0.63 (−0.11 to 1.37) | 0.091 |
| Chronic kidney disease, renin nuclei, % of vascular mural nuclei, unadjusted | 72 | 6.86 (1.41 to 12.30) | 0.014 |
| Chronic kidney disease, renin nuclei, % of vascular mural nuclei, adjusted | 72 | 7.13 (1.38 to 12.88) | 0.016 |

*Supplementary Table S4, continued: D. The 50 renin-cell genes with the smallest P (treated against untreated, all diagnoses) in KPMP*

| Data and model | Participants | Same direction, *n* of *N* (%) | Binomial *P* |
|---|---|---|---|
| KPMP renin nuclei, unadjusted | 40 | 36 of 48 (75%) | <0.001 |
| KPMP renin nuclei, adjusted | 40 | 29 of 48 (60%) | 0.193 |

*E. Tissue measures per NPX of plasma renin, IgA nephropathy, with RAS blockade in the model*

| Measure | Patients | Estimate (95% CI) | *P* |
|---|---|---|---|
| Renin cells per glomerulus | 50 | 1.23 (0.13 to 2.34) | 0.029 |
| Renin share of mural cells at the vascular pole, % | 50 | 8.14 (3.59 to 12.69) | <0.001 |
| Macula densa cells per glomerulus | 50 | 0.16 (−0.14 to 0.45) | 0.286 |
| Pole smooth muscle cells per glomerulus | 50 | −0.27 (−1.30 to 0.76) | 0.595 |
| Distant renin arterioles per $mm^2$ | 46 | 0.03 (−0.05 to 0.10) | 0.465 |

Patients with IgA nephropathy, 59 (26 treated), unless stated; basic and adjusted models as in Supplementary Table S1. A, the stricter renin-cell definition counts mural-lineage cells with at least three nuclear *REN* transcripts whatever their label; batch models without the batch term (batch 1, 7 of 24 patients treated; batch 2, 19 of 35). B, negative binomial regression with the number of glomeruli as exposure; batch rate ratios pooled by inverse variance. C, KPMP open-access data (adjusted, age-bin midpoint, sex and, for all participants, enrollment category). D, genes measured in both data sets; binomial *P* against 50%. E, least squares of each measure on plasma renin with age, sex, sequencing batch and RAS blockade; patients with plasma within 7 days of biopsy. IgAN, IgA nephropathy; KPMP, Kidney Precision Medicine Project; NPX, normalized protein expression; RAS, renin–angiotensin system.

#### Supplementary Table S5. Renin cells against arteriolar smooth muscle

*A. The 12 genes with the largest moderated t (higher in renin cells)*

| Gene | $log_2$ FC (95% CI) | Moderated *t* |
|---|---|---|
| *REN* | 9.44 (9.17 to 9.71) | 70.5 |
| *TRPM3* | 3.51 (3.25 to 3.77) | 27.5 |
| *KCNJ2* | 3.02 (2.78 to 3.25) | 25.7 |
| *MECOM* | 3.09 (2.82 to 3.36) | 23.2 |
| *ADORA1* | 3.36 (3.06 to 3.65) | 22.8 |
| *ABCC8* | 2.35 (2.08 to 2.62) | 17.6 |
| *CD55* | 2.24 (1.98 to 2.50) | 17.3 |
| *POSTN* | 2.47 (2.15 to 2.79) | 15.4 |
| *GATA3* | 2.14 (1.85 to 2.42) | 15.2 |
| *GJA5* | 2.02 (1.75 to 2.29) | 14.9 |
| *PPFIA4* | 1.45 (1.23 to 1.67) | 13.1 |
| *HIF1A* | 1.36 (1.14 to 1.58) | 12.6 |

*B. The 12 genes with the smallest moderated t (lower in renin cells)*

| Gene | $log_2$ FC (95% CI) | Moderated *t* |
|---|---|---|
| *ACTA2* | −2.69 (−2.90 to −2.49) | −26.4 |
| *VIM* | −1.83 (−2.00 to −1.66) | −21.6 |
| *AOC3* | −2.39 (−2.63 to −2.14) | −19.7 |
| *JAG1* | −1.83 (−2.02 to −1.63) | −18.8 |
| *MCAM* | −1.52 (−1.70 to −1.35) | −17.4 |
| *FLNA* | −1.61 (−1.80 to −1.42) | −17.3 |
| *PLN* | −1.79 (−2.00 to −1.57) | −16.9 |
| *COL1A1* | −2.19 (−2.46 to −1.92) | −16.4 |
| *ENTPD1* | −1.96 (−2.21 to −1.72) | −16.2 |
| *MAP3K7CL* | −1.85 (−2.09 to −1.61) | −15.3 |
| *VEGFA* | −1.69 (−1.93 to −1.46) | −14.4 |
| *LDB3* | −1.93 (−2.20 to −1.66) | −14.2 |

Nuclear transcripts summed per patient over all renin cells of the section, against the arteriolar smooth muscle of the same patient (all diagnoses, 109 patients with at least 10 renin cells), limma-voom with patient blocks and transcript yield; 342 genes tested, 75 higher and 123 lower in renin cells at $q < 0.05$; every gene shown has $q < 0.001$. Every gene tested is in Supplementary Data 1. FC, fold change.

#### Supplementary Table S6. Renin cells of treated against untreated patients

*A. Genes of renin cells*

| | All diagnoses (84; 158 genes) | | | IgAN (47; 145) | | | Non-diabetic glomerular disease (69; 139) | | | KPMP |
|---|---|---|---|---|---|---|---|---|---|---|
| Gene | $\log_2$ FC (95% CI) | *P* | *q* | $\log_2$ FC | *P* | *q* | $\log_2$ FC | *P* | *q* | $\log_2$ FC |
| *REN* | 0.51 (0.18 to 0.84) | 0.003 | 0.216 | 0.52 | 0.016 | 0.396 | 0.52 | 0.005 | 0.225 | 1.11 |
| *TRPM3* | 0.52 (0.18 to 0.86) | 0.003 | 0.216 | 0.49 | 0.042 | 0.484 | 0.55 | 0.005 | 0.225 | 0.43 |
| *PRKAR1A* | 0.56 (0.17 to 0.95) | 0.005 | 0.258 | 0.68 | 0.012 | 0.355 | 0.69 | 0.001 | 0.208 | 0.19 |
| *ABCC8* | 0.66 (0.18 to 1.15) | 0.007 | 0.259 | 0.23 | 0.460 | 0.897 | 0.46 | 0.072 | 0.524 | 0.64 |
| *LBH* | −0.59 (−1.03 to −0.14) | 0.009 | 0.259 | −0.64 | 0.008 | 0.355 | −0.43 | 0.056 | 0.524 | 0.30 |
| *PDGFC* | 0.62 (0.15 to 1.09) | 0.010 | 0.259 | 0.62 | 0.043 | 0.484 | 0.58 | 0.026 | 0.453 | −0.12 |
| *CBWD2* | 0.52 (0.10 to 0.94) | 0.014 | 0.306 | – | – | – | – | – | – | 0.06 |
| *TINAGL1* | 0.37 (0.06 to 0.68) | 0.017 | 0.306 | 0.43 | 0.045 | 0.484 | 0.38 | 0.021 | 0.453 | 0.37 |
| *NT5DC2* | 0.48 (0.08 to 0.88) | 0.019 | 0.306 | 0.37 | 0.156 | 0.648 | 0.34 | 0.123 | 0.590 | 0.31 |
| *SOX2-OT* | 0.62 (0.10 to 1.14) | 0.019 | 0.306 | 0.66 | 0.030 | 0.484 | 0.78 | 0.006 | 0.225 | 0.65 |
| *C4A* | −0.58 (−1.09 to −0.08) | 0.023 | 0.317 | −0.47 | 0.104 | 0.580 | −0.62 | 0.027 | 0.453 | – |
| *PHLDB2* | −0.51 (−0.96 to −0.06) | 0.024 | 0.317 | −0.64 | 0.012 | 0.355 | −0.51 | 0.029 | 0.453 | 0.27 |
| *RBPMS* | 0.43 (−0.02 to 0.88) | 0.057 | 0.608 | 0.85 | 0.002 | 0.355 | 0.45 | 0.067 | 0.524 | 0.63 |
| *ENG* | 0.42 (−0.02 to 0.86) | 0.061 | 0.608 | 0.75 | 0.007 | 0.355 | 0.53 | 0.036 | 0.498 | 0.55 |
| *CALM3* | −0.41 (−0.85 to 0.03) | 0.065 | 0.608 | −0.06 | 0.842 | 0.952 | −0.25 | 0.289 | 0.802 | −0.50 |
| *TGM2* | 0.46 (−0.04 to 0.95) | 0.066 | 0.608 | – | – | – | – | – | – | 0.24 |
| *RAC1* | 0.38 (−0.04 to 0.79) | 0.072 | 0.608 | 0.37 | 0.183 | 0.704 | 0.43 | 0.064 | 0.524 | −0.68 |
| *POGZ* | −0.40 (−0.85 to 0.05) | 0.076 | 0.608 | −0.57 | 0.051 | 0.484 | −0.37 | 0.132 | 0.594 | 0.57 |
| *GTF2I* | −0.40 (−0.85 to 0.05) | 0.076 | 0.608 | −0.24 | 0.396 | 0.808 | −0.42 | 0.097 | 0.541 | 0.06 |
| *EPAS1* | 0.32 (−0.05 to 0.69) | 0.081 | 0.608 | 0.46 | 0.053 | 0.484 | 0.38 | 0.058 | 0.524 | 0.46 |
| *ALG11* | 0.45 (−0.06 to 0.96) | 0.083 | 0.608 | 0.65 | 0.035 | 0.484 | 0.56 | 0.048 | 0.524 | 0.28 |
| *PTP4A3* | 0.41 (−0.06 to 0.87) | 0.085 | 0.608 | 0.48 | 0.070 | 0.484 | 0.54 | 0.027 | 0.453 | 0.45 |
| *VIM* | −0.30 (−0.66 to 0.05) | 0.090 | 0.612 | −0.29 | 0.221 | 0.714 | −0.28 | 0.158 | 0.667 | −0.65 |
| *SFPQ* | 0.32 (−0.06 to 0.70) | 0.093 | 0.612 | 0.36 | 0.170 | 0.685 | 0.36 | 0.080 | 0.524 | 0.02 |
| *DDX39B* | 0.32 (−0.07 to 0.72) | 0.100 | 0.630 | 0.27 | 0.287 | 0.769 | 0.27 | 0.200 | 0.730 | 0.44 |

*Supplementary Table S6, continued: B. Cell-level score of the renin-cell program*

| Comparison | Patients | Difference (95% CI) | *P* |
|---|---|---|---|
| Score with all weighted genes | | | |
| Treated minus untreated, IgAN, basic | 54 | 0.02 (−0.08 to 0.11) | 0.659 |
| Treated minus untreated, IgAN, adjusted | 54 | 0.02 (−0.09 to 0.14) | 0.668 |
| Treated minus untreated, non-diabetic glomerular disease, basic | 79 | 0.04 (−0.03 to 0.11) | 0.248 |
| Per step of nuclear *REN* count (1, 2–4, ≥5), same patient | 126 | 0.20 (0.18 to 0.23) | <0.001 |
| Score without *REN* | | | |
| Treated minus untreated, IgAN, basic | 54 | 0.04 (−0.05 to 0.13) | 0.312 |
| Distant renin arteriole minus vascular pole, same patient | 26 | −0.37 (−0.61 to −0.12) | 0.004 |
| Distant renin arteriole minus vascular pole, same patient, IgAN | 14 | −0.45 (−0.78 to −0.12) | 0.011 |
| Other locations minus vascular pole, same patient | 107 | −0.18 (−0.24 to −0.12) | <0.001 |
| Per step of nuclear *REN* count (1, 2–4, ≥5), same patient | 126 | 0.06 (0.02 to 0.09) | <0.001 |

A, nuclear transcripts summed per patient over renin cells (at least 10 per patient), treated minus untreated, basic model with transcript yield (all diagnoses with diagnosis terms); in the group heads, patients and genes tested. The 25 genes with the smallest *P* in all diagnoses, their estimates in IgA nephropathy and in non-diabetic glomerular disease, and the KPMP $\log_2$ fold change of renin nuclei by RAS blockade (KPMP field, 69 on and 47 not on treatment); –, no estimate. Every gene is in Supplementary Data 1. B, cell-level score of the renin-cell program: the log-normalized nuclear counts of each cell projected on the $\log_2$ fold changes of the genes at $q < 0.05$ between renin cells and arteriolar smooth muscle, scaled so that the average arteriolar smooth muscle cell scores 0 and the average renin cell 1; patient medians of renin cells (patients with at least five renin cells; locations and transcript counts, at least three cells each), basic and adjusted models as in Supplementary Table S1, locations and transcript counts within patients with fixed effects and errors clustered by patient. FC, fold change; IgAN, IgA nephropathy; KPMP, Kidney Precision Medicine Project; RAS, renin–angiotensin system.

#### Supplementary Table S7. Mouse renin-cell gene sets and profiles in renin cells of treated patients

*A. Gene sets*

| Gene set (mouse data) | Genes | IgAN, treated vs untreated, basic (47): Estimate (null range) | IgAN, treated vs untreated, basic (47): *P*, camera | IgAN, adjusted: Estimate (null range) | IgAN, adjusted: *P*, camera | Renin cells vs smooth muscle: Estimate (null range) | Renin cells vs smooth muscle: *P*, camera |
|---|---|---|---|---|---|---|---|
| Matrix-secretory, all genes (renin deletion) | 13; 29 | 0.48 (−0.64 to 0.67) | 0.230 | 0.34 (−0.58 to 0.60) | 0.356 | −3.17 (−2.53 to 2.61) | 0.063 |
| Matrix-secretory, secreted proteins[a] (renin deletion) | 5; 10 | 1.31 (−0.98 to 1.07) | 0.037 | 1.16 (−0.87 to 0.97) | 0.041 | −4.45 (−4.11 to 4.40) | 0.083 |
| Endocrine identity (renin deletion) | 3; 6 | −0.80 (−1.23 to 1.39) | 0.233 | −0.21 (−1.10 to 1.23) | 0.716 | 2.68 (−5.38 to 5.65) | 0.380 |
| Higher after captopril (captopril, low salt, 2 weeks) | 12; 19 | −0.12 (−0.67 to 0.70) | 0.781 | −0.30 (−0.60 to 0.61) | 0.479 | −6.60 (−3.06 to 3.21) | 0.006 |

*B. Profiles*

| Gene set or profile (mouse data) | Genes | IgAN, treated vs untreated, basic (47): Estimate (null range) | IgAN, treated vs untreated, basic (47): *P* | IgAN, adjusted: Estimate (null range) | IgAN, adjusted: *P* | Renin cells vs smooth muscle: Estimate (null range) | Renin cells vs smooth muscle: *P* |
|---|---|---|---|---|---|---|---|
| Renin deletion minus wild type (renin deletion) | 123; 303 | 0.09 (−0.18 to 0.17) | 0.306 | 0.04 (−0.18 to 0.18) | 0.690 | −0.12 (−0.11 to 0.11) | 0.039 |
| Captopril minus untreated (captopril, low salt, 2 weeks) | 127; 300 | −0.04 (−0.18 to 0.17) | 0.615 | −0.02 (−0.17 to 0.18) | 0.831 | −0.20 (−0.11 to 0.11) | <0.001 |
| Recruited minus sham renin cells (captopril, low salt, 1 week) | 126; 305 | −0.05 (−0.18 to 0.17) | 0.581 | −0.07 (−0.17 to 0.17) | 0.407 | −0.06 (−0.11 to 0.12) | 0.327 |
| Low minus high perfusion pressure (aortic coarctation) | 126; 305 | 0.07 (−0.17 to 0.17) | 0.460 | 0.04 (−0.18 to 0.18) | 0.664 | 0.06 (−0.11 to 0.11) | 0.284 |

Mouse renin-cell gene sets and profiles against the renin-cell differences of this study; genes, the measured genes in the comparison of treated with untreated patients; then in that of renin cells with smooth muscle. Gene sets, mean moderated *t* of the measured set genes minus that of all other measured genes; profiles, Spearman correlation of the mouse and human differences over the genes measured in both. Null range, 2.5th to 97.5th percentiles of 10,000 random gene sets of the same size (gene-label permutations for profiles); *P*, two-sided permutation (profiles) or camera, a competitive test with the inter-gene correlation estimated from the data (gene sets; limma camera on the same fits; random-set *P* in Supplementary Data 1). Human differences, renin cells of treated against untreated patients (at least 10 renin cells; patients in the group heads) and renin cells against arteriolar smooth muscle of the same patients. Mouse genes were mapped to one-to-one human orthologs (Ensembl). Sources: renin deletion, GSE180873 and the published differential-expression tables; captopril (with a low-salt diet for 2 weeks), GSE42712 against GSE17138; captopril with a low-salt diet for 1 week, GSE146386 against the sham-operated mice of GSE157699; low perfusion pressure, GSE157699. Gene sets with fewer than three measured genes were not tested. Estimates over all diagnoses are in Supplementary Data 1. [a]The 5 secreted-protein genes measured in IgA nephropathy, treated against untreated (basic model), $\log_2$ fold change: *APP* 0.45 (*P* = 0.093), *BGN* 0.57 (*P* = 0.020), *COL18A1* 0.27 (*P* = 0.231), *FN1* 0.60 (*P* = 0.064) and *SOD3* −0.20 (*P* = 0.470). IgAN, IgA nephropathy.

#### Supplementary Table S8. Macula densa markers, macula densa expression by treatment and ligand–receptor pairs

*A. Macula densa against cortical thick ascending limb, all diagnoses (49 patients, 1,446 genes tested)*

| Gene | $\log_2$ FC (95% CI) | *P* | *q* |
|---|---|---|---|
| *ROBO2* | 2.52 (2.18 to 2.87) | <0.001 | <0.001 |
| *MEST* | 1.86 (1.54 to 2.18) | <0.001 | <0.001 |
| *DDX3Y* | 2.15 (1.70 to 2.59) | <0.001 | <0.001 |
| *CALCR* | 1.37 (1.03 to 1.70) | <0.001 | <0.001 |
| *STIM2* | 1.27 (0.95 to 1.59) | <0.001 | <0.001 |
| *TNFRSF11B* | 1.38 (1.03 to 1.72) | <0.001 | <0.001 |
| *PAPPA2* junction probe (ENST00000367662) | 1.30 (0.95 to 1.66) | <0.001 | <0.001 |
| *MYH10* | 1.06 (0.74 to 1.37) | <0.001 | <0.001 |
| *DUSP6* | 1.14 (0.80 to 1.48) | <0.001 | <0.001 |
| *UMOD* | −2.75 (−3.07 to −2.44) | <0.001 | <0.001 |
| *EGF* | −2.59 (−2.89 to −2.29) | <0.001 | <0.001 |
| *CXCL14* | −2.61 (−2.94 to −2.29) | <0.001 | <0.001 |
| *CXCL12* | −2.42 (−2.73 to −2.12) | <0.001 | <0.001 |
| *TMEM52B* | −2.38 (−2.69 to −2.08) | <0.001 | <0.001 |
| *NOS1*[a] | 0.62 (0.28 to 0.96) | <0.001 | 0.001 |
| *PAPPA2*[a] | 0.43 (0.08 to 0.78) | 0.014 | 0.036 |

*B. Macula densa, IgA nephropathy, treated against untreated (21 patients, 98 genes tested)*

| Gene | $\log_2$ FC (95% CI) | *P* | *q* |
|---|---|---|---|
| *MUC1* | 1.37 (0.32 to 2.43) | 0.011 | 0.581 |
| *COL4A3* | 1.18 (0.26 to 2.10) | 0.012 | 0.581 |
| *DAP* | −1.55 (−2.86 to −0.25) | 0.020 | 0.581 |
| *GPRC5C* | −1.44 (−2.75 to −0.12) | 0.032 | 0.581 |
| *HNRNPM* | 1.17 (−0.03 to 2.38) | 0.055 | 0.581 |
| *CGNL1* | 1.18 (−0.04 to 2.40) | 0.057 | 0.581 |
| *UMOD*[b] | −0.74 (−1.55 to 0.07) | 0.073 | 0.581 |

*Supplementary Table S8, continued: C. TAL within 15 µm of a glomerulus, IgA nephropathy, treated against untreated (35 patients, 317 genes tested)*

| Gene | $\log_2$ FC (95% CI) | *P* | *q* |
|---|---|---|---|
| *SLC5A3* | −0.92 (−1.59 to −0.26) | 0.007 | 0.593 |
| *OSGIN1* | 0.93 (0.24 to 1.63) | 0.009 | 0.593 |
| *UMOD* | −0.71 (−1.29 to −0.12) | 0.018 | 0.593 |
| *CA12* | −0.52 (−0.95 to −0.08) | 0.019 | 0.593 |
| *GADD45A* | −0.77 (−1.44 to −0.11) | 0.022 | 0.593 |
| *PTGER3* | −0.87 (−1.62 to −0.12) | 0.024 | 0.593 |
| *EGF*[b] | −0.92 (−1.78 to −0.05) | 0.037 | 0.593 |

*D. Ligand–receptor pairs between macula densa and renin cells, IgA nephropathy, treated minus untreated*

| Ligand–receptor | Difference (95% CI) | *P* |
|---|---|---|
| *FGF9–FGFR1* | −0.001 (−0.003 to 0.001) | 0.194 |
| *CD46–JAG1* | 0.000 (−0.003 to 0.002) | 0.696 |
| *SDC2–ADGRA2* | −0.004 (−0.010 to 0.002) | 0.134 |
| PGE2 (*PTGES3*)–*PTGER2* | 0.013 (−0.013 to 0.039) | 0.319 |

Macula densa, thick ascending limb cells within 15 µm of a glomerulus and within 40 µm of its vascular pole (the centroid of the smooth muscle cells and juxtaglomerular-labeled cells without a nuclear *REN* transcript within 20 µm). A, nuclear transcripts summed per patient over macula densa cells against cortical thick ascending limb cells of the same patient (more than 50 and up to 300 µm from a glomerulus), patients with at least 10 macula densa cells; the 9 genes with the largest and 5 with the smallest moderated *t* and the literature genes *NOS1*, *PAPPA2* and *ROBO2* ([a]added outside the ranking; no estimate for *PTGS2*); a junction probe targets one transcript isoform. B, C, the six genes with the smallest *P* ([b]genes named in the text, added outside the ranking), basic model with transcript yield; B, patients with at least 10 macula densa cells; of the 59 genes of A with $q < 0.05$ and at least a twofold difference that were tested in C, 6 were higher and 53 lower in the macula densa; 4 of the 6 were higher and 41 of the 53 lower in treated patients (adjusted model, 4 and 37). D, CellChatDB v2 (human): 3,233 interactions, 1,109 with every ligand and receptor gene on the panel; no pair had both a ligand higher in macula densa than in cortical thick ascending limb and a receptor higher in renin cells than in smooth muscle ($q < 0.1$). Shown, pairs with every ligand gene detected in at least 5% of macula densa cells and every receptor gene in at least 5% of pole renin cells (ligand, then receptor genes); difference in the pole score (mean ligand expression of macula densa cells times mean receptor expression of renin cells at the same pole), basic model with errors clustered by patient; a metabolite ligand is named with its synthesizing enzyme in parentheses. FC, fold change; TAL, thick ascending limb.

**Supplementary Table S9. Thick ascending limb genes at the vascular pole and renin cells at the same pole**

*Thick ascending limb cells within 15 µm of the glomerulus, all diagnoses*

| | | | | Interaction *P* | |
|---|---|---|---|---|---|
| Gene | $\log_2$ FC per doubling (95% CI) | *P* | *q* | All diagnoses | IgA nephropathy |
| *MEST* | 0.25 (0.18 to 0.32) | <0.001 | <0.001 | 0.305 | 0.106 |
| *ACSL4* | 0.23 (0.15 to 0.31) | <0.001 | <0.001 | 0.644 | 0.485 |
| *COL4A3* | 0.18 (0.10 to 0.26) | <0.001 | <0.001 | 0.703 | 0.082 |
| *IRX1* | 0.19 (0.10 to 0.27) | <0.001 | <0.001 | 0.188 | 0.613 |
| *LGR4* | 0.15 (0.08 to 0.22) | <0.001 | <0.001 | 0.094 | 0.052 |
| *SERPINA5* | 0.17 (0.09 to 0.26) | <0.001 | 0.005 | 0.716 | 0.729 |
| *CA12* | 0.16 (0.08 to 0.25) | <0.001 | 0.005 | 0.758 | 0.226 |
| *HSPA8* | 0.13 (0.06 to 0.20) | <0.001 | 0.006 | 0.412 | 0.197 |
| *MYH10* | 0.12 (0.05 to 0.19) | <0.001 | 0.018 | 0.751 | 0.697 |
| *CA4* | 0.12 (0.05 to 0.19) | <0.001 | 0.019 | 0.830 | 0.566 |
| *ROBO2* | 0.12 (0.05 to 0.18) | <0.001 | 0.019 | 0.423 | 0.594 |
| *TNFRSF11B* | 0.11 (0.05 to 0.18) | <0.001 | 0.027 | 0.553 | 0.143 |
| *VDR* | 0.11 (0.04 to 0.18) | <0.001 | 0.033 | 0.041 | 0.616 |
| *KCNJ16* | 0.13 (0.05 to 0.21) | <0.001 | 0.039 | 0.581 | 0.350 |
| *GBE1* | 0.09 (0.03 to 0.15) | 0.001 | 0.063 | 0.104 | 0.126 |
| *SQSTM1* | 0.10 (0.03 to 0.16) | 0.001 | 0.063 | 0.995 | 0.983 |
| *IVNS1ABP* | 0.12 (0.04 to 0.20) | 0.001 | 0.064 | 0.248 | 0.175 |
| *MAN1A1* | 0.11 (0.04 to 0.18) | 0.001 | 0.064 | 0.643 | 0.692 |
| *HNF1B* | 0.10 (0.03 to 0.17) | 0.001 | 0.064 | 0.779 | 0.664 |
| *ENOX1* | 0.09 (0.03 to 0.16) | 0.001 | 0.065 | 0.452 | 0.973 |
| *CLCN5* | 0.11 (0.04 to 0.19) | 0.002 | 0.065 | 0.509 | 0.808 |
| *ATP1A1* | 0.13 (0.04 to 0.21) | 0.002 | 0.065 | 0.866 | 0.890 |
| *CTSL* | 0.09 (0.03 to 0.15) | 0.002 | 0.065 | 0.507 | 0.885 |
| *POLR2B* | −0.08 (−0.13 to −0.02) | 0.002 | 0.078 | 0.664 | 0.719 |
| *COBLL1* | 0.10 (0.03 to 0.17) | 0.002 | 0.088 | 0.322 | 0.363 |
| *KCNJ1* | 0.10 (0.03 to 0.17) | 0.003 | 0.090 | 0.820 | 0.790 |
| *MYO6* | 0.10 (0.03 to 0.17) | 0.003 | 0.090 | 0.179 | 0.125 |
| *DHX30* | −0.07 (−0.13 to −0.02) | 0.003 | 0.090 | 0.457 | – |
| *RBBP8* | 0.09 (0.03 to 0.16) | 0.003 | 0.093 | 0.772 | 0.863 |
| *PCK1* | 0.09 (0.02 to 0.15) | 0.003 | 0.093 | 0.846 | 0.897 |

Nuclear transcripts of thick ascending limb cells within 15 µm of each glomerulus summed per vascular pole (poles with at least three such cells, 537 poles of 104 participants with at least two poles), limma-voom with participant blocks and transcript yield as a covariate; exposure, $\log_2(1 + \text{renin cells within 20 µm of the same glomerulus})$. Genes with at least 20 counts detected in at least 20% of the poles (925 genes); shown, the 30 genes at $q < 0.1$. Interaction *P*, the slope in treated minus untreated patients (patients with kidney disease and a medication record: all diagnoses 74, IgA nephropathy 44 patients). Estimates in treated and untreated patients are in Supplementary Data 1. Of the genes of Supplementary Table S8 A with $q < 0.05$ and at least a twofold difference that were tested here, 11 of the 12 higher in the macula densa had a positive slope and 28 of the 60 lower in the macula densa a negative slope. FC, fold change.

**Supplementary Table S10. Juxtaglomerular measures, the clinical state at biopsy and the kidney course in IgA nephropathy**

*A. Clinical state at biopsy*

| Measure, per SD | Patients | eGFR, mL/min/1.73 m$^2$ | log UPCR | Mean arterial pressure, mmHg |
|---|---|---|---|---|
| Renin cells per glomerulus, $\log_2(1 + x)$ | 59 | 0.71 (−4.35 to 5.76) | 0.00 (−0.37 to 0.36) | −1.92 (−5.01 to 1.16) |
| Renin share of pole mural cells | 59 | 1.23 (−3.85 to 6.32) | −0.12 (−0.49 to 0.24) | −1.66 (−5.04 to 1.72) |
| Labeled pole cells with nuclear *REN* | 56 | 1.19 (−4.55 to 6.93) | 0.21 (−0.10 to 0.53) | −0.87 (−4.73 to 2.99) |
| Macula densa cells per glomerulus | 59 | 3.01 (−0.93 to 6.96) | −0.29 (−0.52 to −0.07) | −1.71 (−4.14 to 0.73) |
| Glomeruli with a macula densa plaque | 59 | 2.50 (−1.38 to 6.38) | −0.24 (−0.50 to 0.02) | −0.78 (−3.71 to 2.16) |
| Plasma renin | 51 | −3.31 (−11.56 to 4.93) | 0.11 (−0.42 to 0.64) | −3.97 (−7.31 to −0.63) |

*B. Kidney course and proteinuria after biopsy*

| Measure, per SD | Followed (events) | Composite outcome, hazard ratio | eGFR slope, mL/min/ 1.73 m$^2$ per year | UPCR at 6 months, $\log_2$ ratio | Partial remission, odds ratio |
|---|---|---|---|---|---|
| Renin cells per glomerulus, $\log_2(1 + x)$ | 57 (13) | 0.82 (0.49 to 1.37) | 0.28 (−0.49 to 1.05) | −0.14 (−0.62 to 0.33) | 1.40 (0.71 to 2.77) |
| Renin share of pole mural cells | 57 (13) | 0.78 (0.45 to 1.36) | 0.24 (−0.52 to 1.00) | −0.13 (−0.57 to 0.32) | 1.37 (0.68 to 2.75) |
| Labeled pole cells with nuclear *REN* | 54 (12) | 0.66 (0.30 to 1.44) | 0.10 (−0.73 to 0.94) | −0.20 (−0.84 to 0.44) | 1.06 (0.52 to 2.15) |
| Macula densa cells per glomerulus | 57 (13) | 0.73 (0.35 to 1.51) | 0.31 (−0.43 to 1.05) | −0.51 (−1.43 to 0.42) | 2.02 (0.94 to 4.32) |
| Glomeruli with a macula densa plaque | 57 (13) | 0.75 (0.29 to 1.89) | 0.25 (−0.50 to 1.00) | −0.49 (−1.96 to 0.98) | 1.53 (0.75 to 3.11) |
| Plasma renin | 49 (12) | 1.06 (0.53 to 2.07) | −0.81 (−1.69 to 0.07) | −0.17 (−0.55 to 0.21) | 1.18 (0.51 to 2.73) |

Estimate (95% CI) per standard deviation of each measure among the patients with it: difference in eGFR, natural log UPCR (g/g), mean arterial pressure, eGFR slope and $\log_2$ of the UPCR at 6 months over the UPCR at biopsy; hazard ratio for the composite outcome; odds ratio for partial remission. Model: age, sex, sequencing batch, RAS blockade, eGFR and log UPCR at biopsy, without the variable that is the outcome of the column; least squares, Cox model, linear mixed model of all eGFR values (measure-by-year term) and logistic regression. Composite outcome, an eGFR at or below 60% of the eGFR at biopsy confirmed at least 28 days later or, at the end of follow-up, without recovery to 80%, or an eGFR below 15 mL/min/1.73 m$^2$ on two occasions (Supplementary Methods); 13 events in 58 patients followed (median follow-up 8.4 years; interquartile range 7.3 to 9.9); the eGFR slope in the same patients. UPCR at 6 months, the value nearest day 183 within days 92 to 273 (57 patients). Partial remission, a UPCR at or below half the value at biopsy and below 1 g/g within 12 months (reached by 38 of 60 patients); rows with fewer patients hold the patients with the measure. Labeled pole cells with nuclear *REN*, the share of juxtaglomerular-labeled cells within 20 µm of a glomerulus that carried a nuclear *REN* transcript; macula densa as in Supplementary Table S1; plasma renin within 7 days of biopsy. Associations with the Oxford classification, estimates within treated and untreated patients and *P* values are in Supplementary Data 1. eGFR, estimated glomerular filtration rate (CKD-EPI 2021); RAS, renin–angiotensin system; UPCR, urine protein-to-creatinine ratio.

**Supplementary Table S11. Juxtaglomerular measures, the clinical state at biopsy and the kidney course in non-diabetic glomerular disease**

*A. Clinical state at biopsy*

| Measure, per SD | Patients | eGFR, mL/min/1.73 $m^2$ | log UPCR | Mean arterial pressure, mmHg |
|---|---|---|---|---|
| Renin cells per glomerulus, $\log_2(1 + x)$ | 89 | −1.03 (−6.17 to 4.11) | 0.05 (−0.17 to 0.27) | −1.70 (−4.60 to 1.20) |
| Renin share of pole mural cells | 88 | −1.65 (−7.81 to 4.52) | −0.02 (−0.24 to 0.20) | −1.81 (−4.95 to 1.33) |
| Labeled pole cells with nuclear *REN* | 82 | −0.18 (−4.95 to 4.59) | 0.16 (−0.06 to 0.37) | −0.98 (−4.18 to 2.23) |
| Macula densa cells per glomerulus | 89 | 2.34 (−0.74 to 5.42) | −0.22 (−0.37 to −0.07) | −1.56 (−3.70 to 0.57) |
| Glomeruli with a macula densa plaque | 89 | 2.33 (−0.96 to 5.62) | −0.18 (−0.35 to −0.02) | −0.81 (−3.20 to 1.58) |
| Plasma renin | 75 | −5.08 (−11.31 to 1.15) | 0.09 (−0.18 to 0.37) | −3.26 (−7.09 to 0.57) |

*B. Kidney course and proteinuria after biopsy*

| Measure, per SD | Followed (events) | Composite outcome, hazard ratio | eGFR slope, mL/min/1.73 $m^2$ per year | UPCR at 6 months, $\log_2$ ratio | Partial remission, odds ratio |
|---|---|---|---|---|---|
| Renin cells per glomerulus, $\log_2(1 + x)$ | 86 (19) | 1.10 (0.68 to 1.76) | −0.25 (−1.75 to 1.24) | −0.20 (−0.69 to 0.30) | 1.32 (0.71 to 2.46) |
| Renin share of pole mural cells | 85 (19) | 0.92 (0.55 to 1.52) | 0.54 (−0.98 to 2.06) | −0.22 (−0.69 to 0.25) | 1.43 (0.73 to 2.77) |
| Labeled pole cells with nuclear *REN* | 79 (17) | 0.71 (0.40 to 1.25) | 0.30 (−1.10 to 1.71) | −0.06 (−0.70 to 0.57) | 0.99 (0.52 to 1.88) |
| Macula densa cells per glomerulus | 86 (19) | 0.77 (0.42 to 1.42) | 0.17 (−1.24 to 1.58) | −0.58 (−1.40 to 0.24) | 1.65 (0.90 to 3.01) |
| Glomeruli with a macula densa plaque | 86 (19) | 0.86 (0.48 to 1.54) | −0.14 (−1.56 to 1.29) | −0.64 (−1.94 to 0.67) | 1.38 (0.77 to 2.48) |
| Plasma renin | 72 (16) | 1.16 (0.68 to 1.96) | 0.14 (−1.66 to 1.94) | −0.25 (−0.65 to 0.16) | 1.12 (0.53 to 2.38) |

Estimate (95% CI) per standard deviation of each measure among the patients with it: difference in eGFR, natural log UPCR (g/g), mean arterial pressure, eGFR slope and $\log_2$ of the UPCR at 6 months over the UPCR at biopsy; hazard ratio for the composite outcome; odds ratio for partial remission. Model: age, sex, sequencing batch, RAS blockade, eGFR and log UPCR at biopsy, without the variable that is the outcome of the column, and diagnosis (Cox models stratified by diagnosis); least squares, Cox model, linear mixed model of all eGFR values (measure-by-year term) and logistic regression; mean arterial pressure recorded for 67 to 80 patients. Composite outcome, an eGFR at or below 60% of the eGFR at biopsy confirmed at least 28 days later or, at the end of follow-up, without recovery to 80%, or an eGFR below 15 mL/min/1.73 $m^2$ on two occasions (Supplementary Methods); 19 events in 87 patients followed (median follow-up 7.8 years; interquartile range 5.6 to 9.2); the eGFR slope in the same patients. UPCR at 6 months, the value nearest day 183 within days 92 to 273 (78 patients). Partial remission, a UPCR at or below half the value at biopsy and below 1 g/g within 12 months (reached by 54 of 82 patients); rows with fewer patients hold the patients with the measure. Labeled pole cells with nuclear *REN*, the share of juxtaglomerular-labeled cells within 20 µm of a glomerulus that carried a nuclear *REN* transcript; macula densa as in Supplementary Table S1; plasma renin within 7 days of biopsy. Estimates within treated and untreated patients and *P* values are in Supplementary Data 1. eGFR, estimated glomerular filtration rate (CKD-EPI 2021); RAS, renin–angiotensin system; UPCR, urine protein-to-creatinine ratio.

#### Supplementary Figures

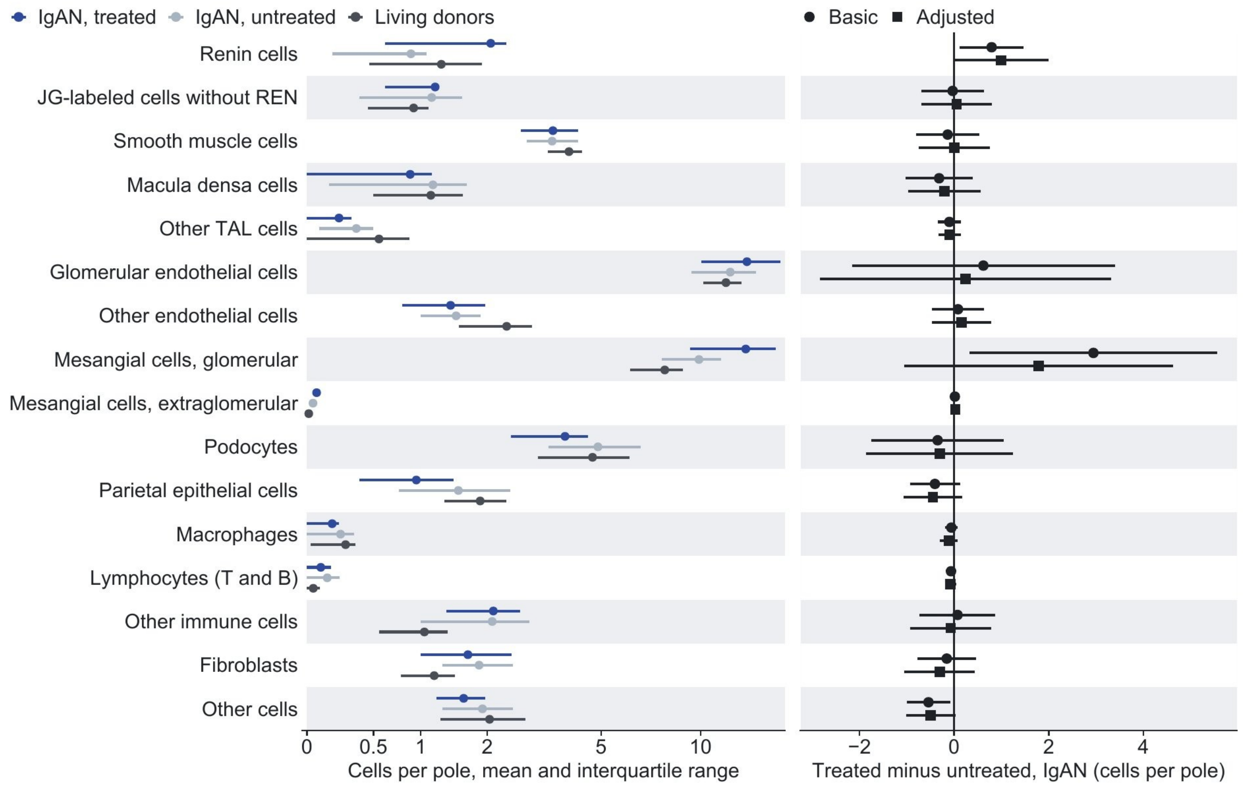


**Supplementary Figure S1. Cells of the vascular pole by treatment.** Cells per pole of each class within 20 µm of the glomerulus and 40 µm of the pole center, by group (mean and interquartile range), and treated minus untreated in IgA nephropathy in the basic and adjusted models (estimates and 95% confidence intervals). IgAN, IgA nephropathy; JG, juxtaglomerular; TAL, thick ascending limb.

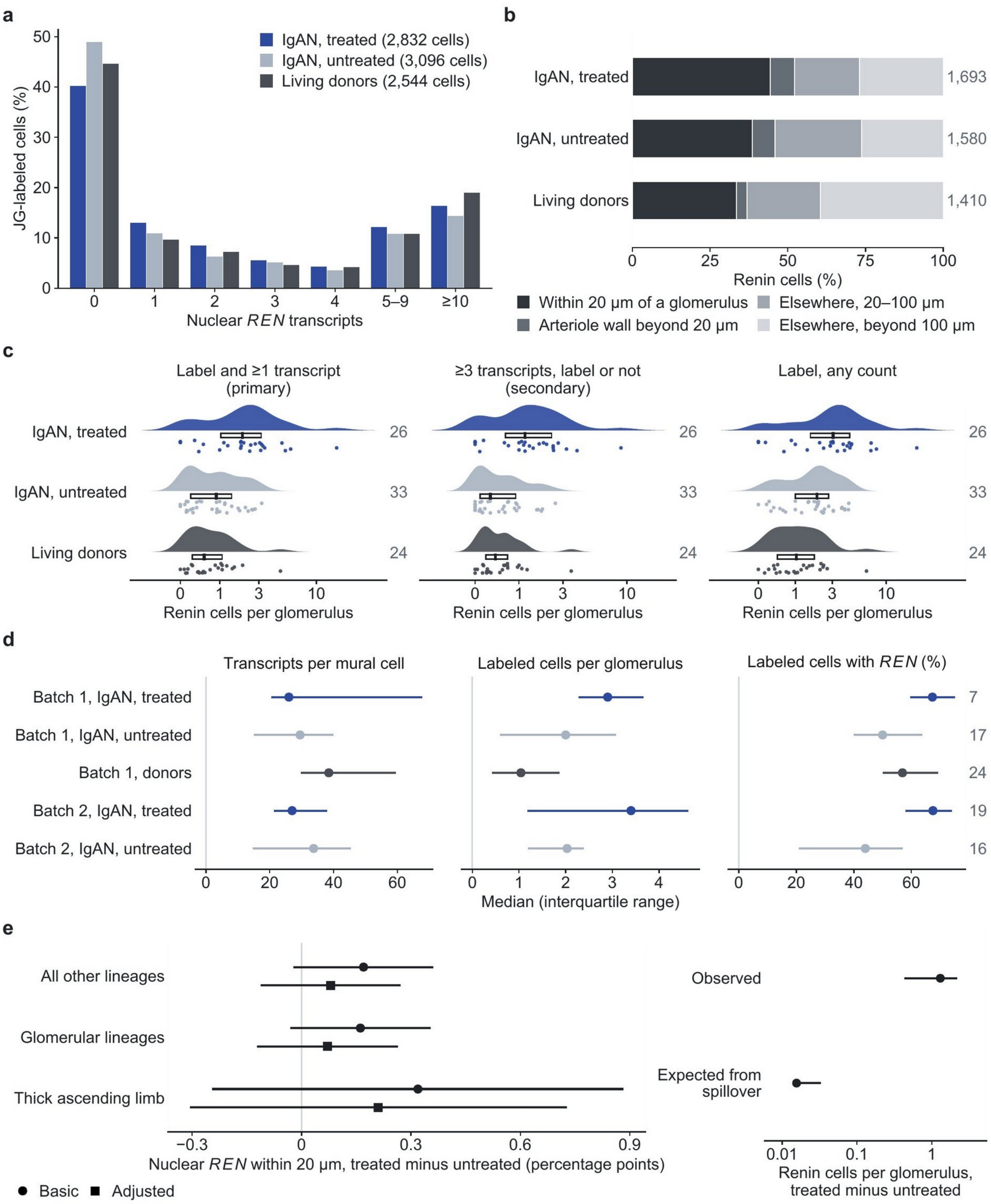


**Supplementary Figure S2. Renin-cell calls and their quality. a**, Cells with the juxtaglomerular (JG) label by nuclear *REN* transcript count, as a percentage of the labeled cells of each group (labeled cells per group in the key). **b**, Location of all renin cells by group: within 20 µm of a glomerulus, in an arteriolar wall beyond 20 µm, elsewhere 20–100 µm and beyond 100 µm from a glomerulus; renin cells per group at right. **c**, Renin cells per glomerulus under three definitions: label and at least one nuclear *REN* transcript (primary), at least three transcripts with or without the label, and label with any count (log(1+x) axis). **d**, Nuclear transcripts per mural cell, labeled cells per glomerulus and labeled cells with a *REN*

transcript by sequencing batch and group: median and interquartile range; patients per row at right. **e**, Left, cells of other lineages within 20 µm of a glomerulus with a nuclear *REN* transcript, treated minus untreated (percentage points; basic and adjusted models); right, the observed treated-minus-untreated difference in renin cells per glomerulus and the difference that spillover at the other-lineage rate would add (logarithmic axis). Rainclouds: density (top), interquartile box with median and participants (points). Points and lines, estimates and 95% confidence intervals. IgAN, IgA nephropathy.

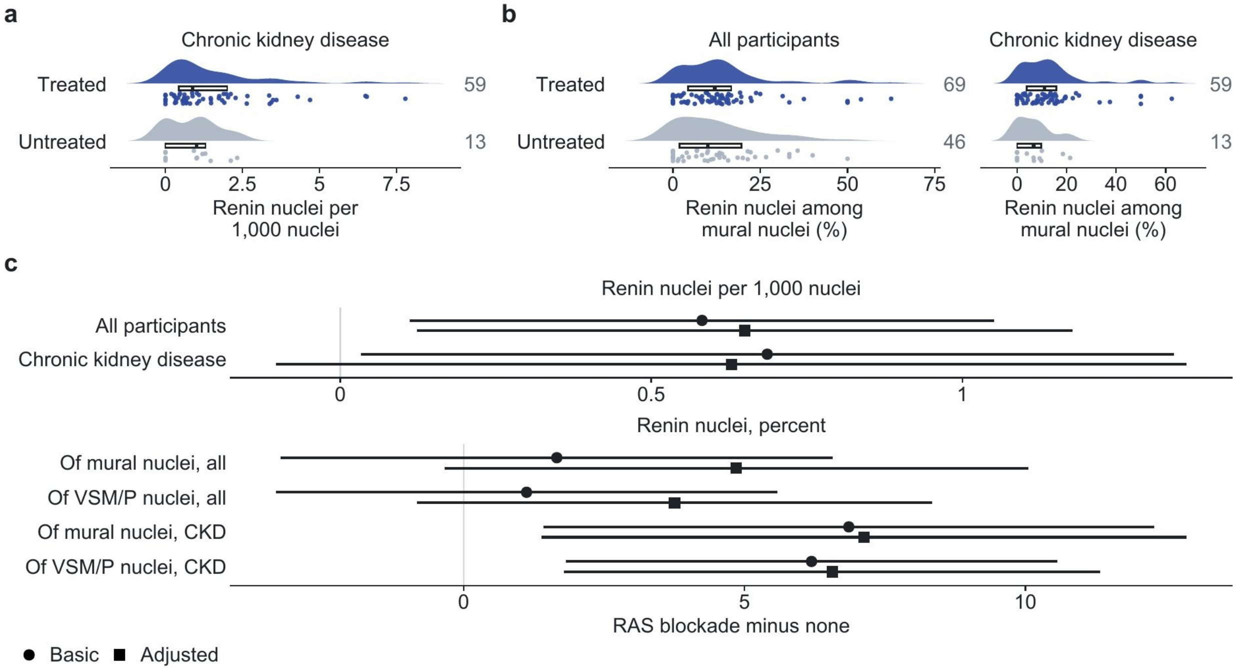


**Supplementary Figure S3. Renin nuclei in the Kidney Precision Medicine Project atlas. a**, Renin nuclei per 1,000 nuclei by RAS blockade in participants with chronic kidney disease (CKD). **b**, Renin nuclei as a percentage of vascular mural nuclei by RAS blockade in all participants and in CKD. **c**, RAS blockade minus none for renin nuclei per 1,000 nuclei and as a percentage of vascular mural nuclei, in all participants and in CKD, unadjusted (basic) and adjusted for age, sex and enrollment category. Rainclouds: density (top), interquartile box with median and participants (points); participants at right. Points and lines, estimates and 95% confidence intervals. KPMP, Kidney Precision Medicine Project; RAS, renin–angiotensin system; VSM/P, vascular smooth muscle cells and pericytes.

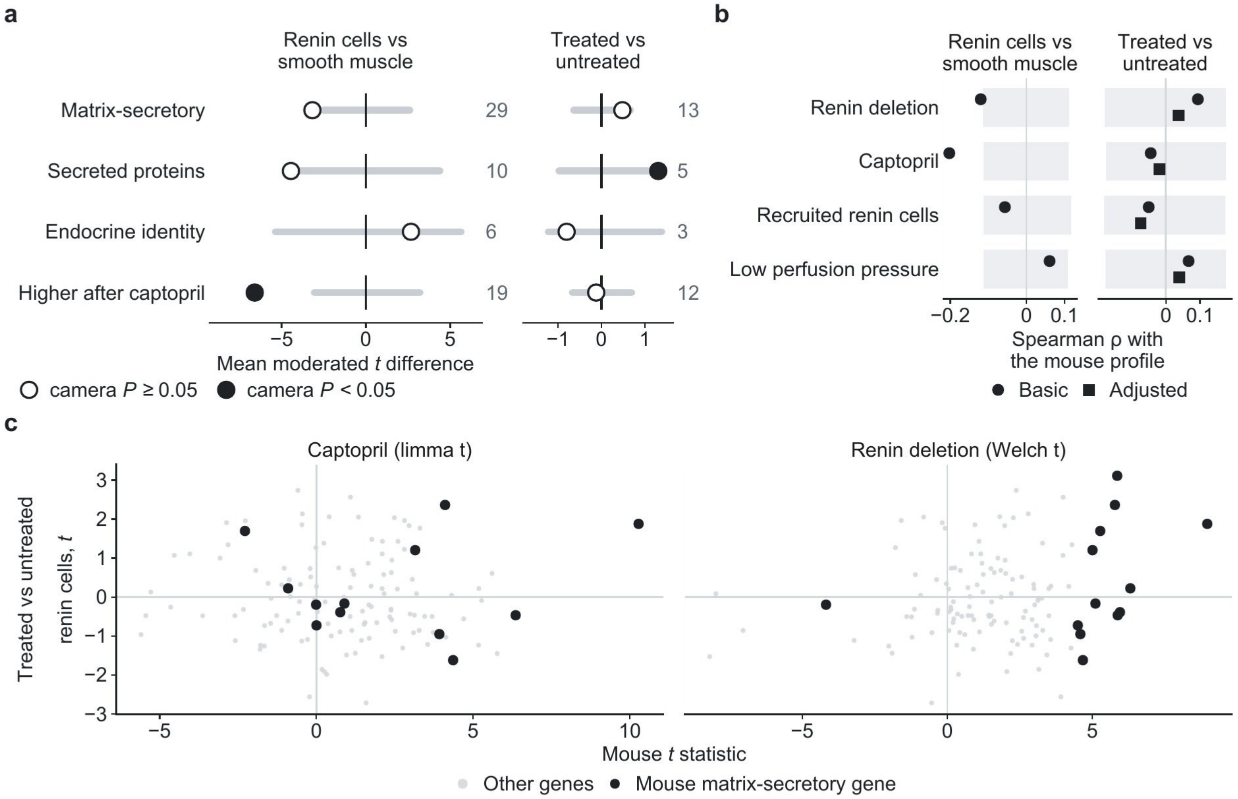


**Supplementary Figure S4. Mouse renin-cell gene sets and profiles. a**, Mouse renin-cell gene sets (matrix-secretory genes after renin deletion and their secreted-protein subset, endocrine identity genes, genes higher after two weeks of captopril) in renin cells against smooth muscle and in treated against untreated patients; gray bars, 95% range of random gene sets of the same size; numbers, genes measured; filled circles, camera $P < 0.05$. **b**, Spearman correlation of the two human contrasts with mouse renin-cell profiles (renin deletion; captopril with a low-salt diet for 2 weeks; recruited renin cells, the same treatment for 1 week; low perfusion pressure), basic and adjusted models; bands, 95% range of random gene sets. **c**, Gene-level comparison with the captopril and renin-deletion profiles (mouse *t* statistic); black, matrix-secretory genes.

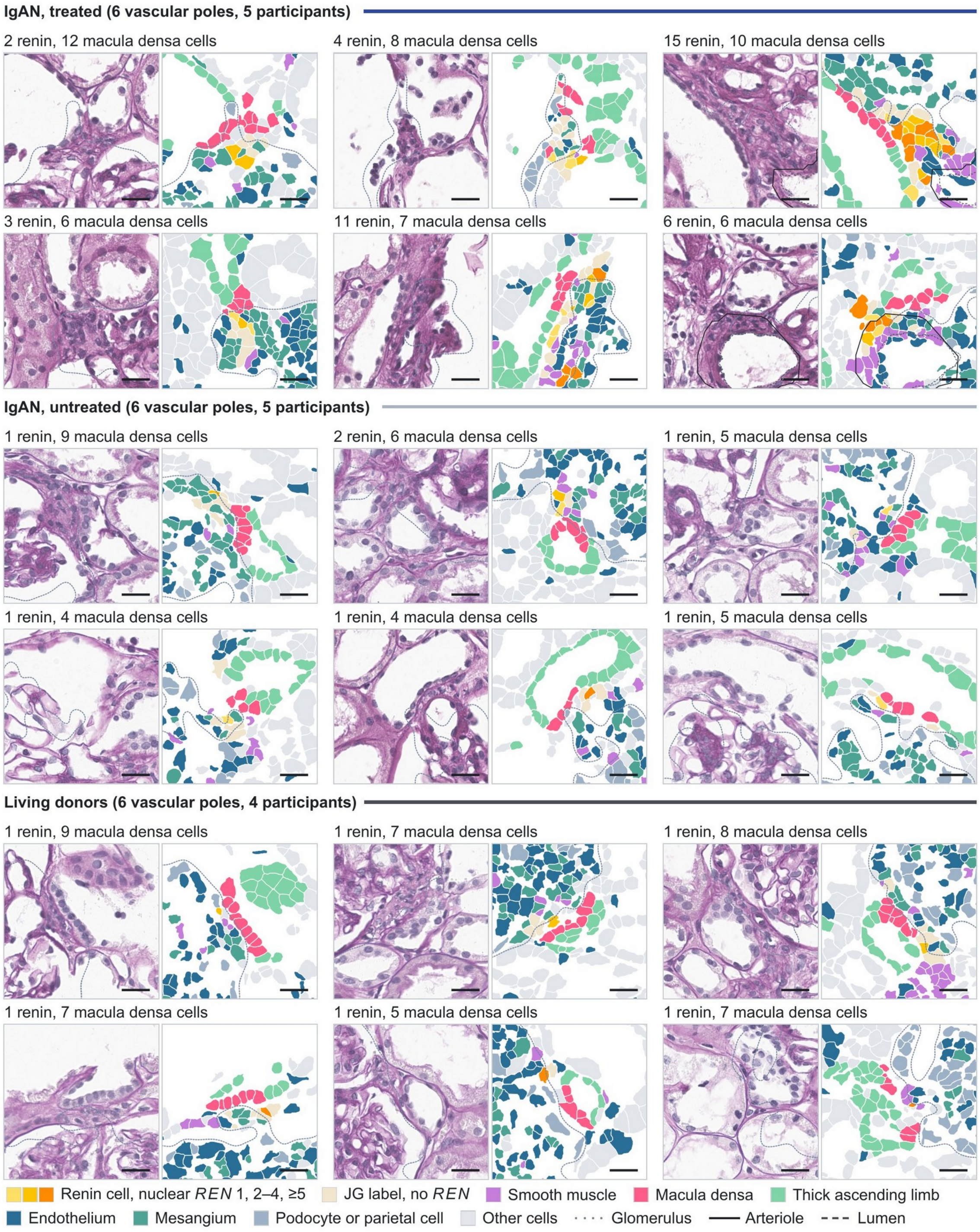


**Supplementary Figure S5. Vascular poles by group.** Six vascular poles per group with a macula densa plaque of at least three cells and a renin cell within 20 µm (110-µm windows), two of each group also shown in Figure 4b; title, renin and macula densa cells. Tissue tiles as in Figure 1. IgAN, IgA nephropathy; JG, juxtaglomerular.

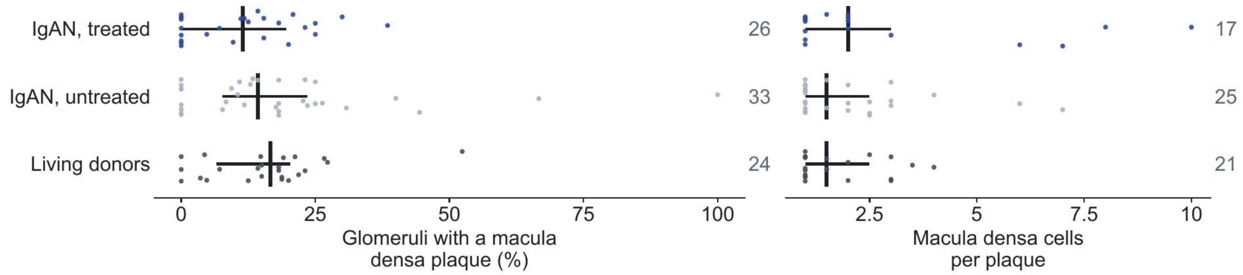


**Supplementary Figure S6. Macula densa plaques.** Glomeruli with a macula densa plaque and macula densa cells per plaque by group (participants with median and interquartile range; participants at right). IgAN, IgA nephropathy.

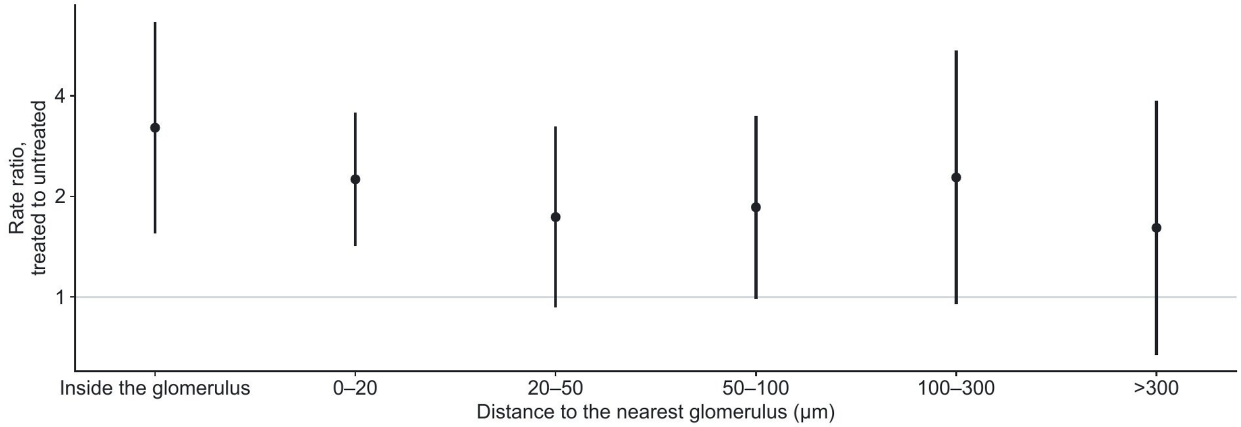


**Supplementary Figure S7. Renin cells by distance from the glomerulus.** Treated-to-untreated rate ratio of renin cells in each band, inside the glomerular outline included, from one negative binomial model over the bands with band tissue area as exposure (logarithmic axis; 59 patients). Points and lines, estimates and 95% confidence intervals.

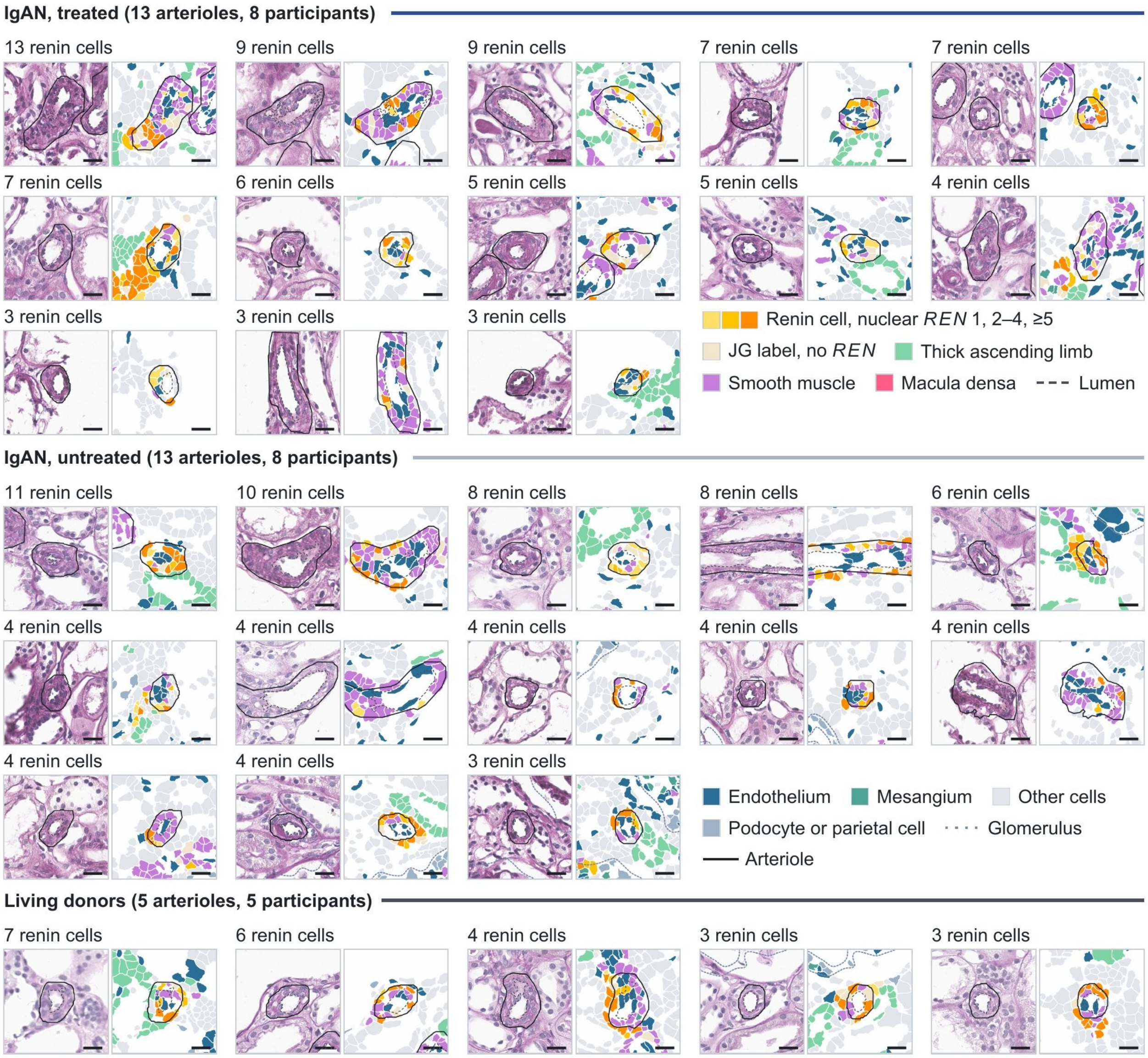


**Supplementary Figure S8. Distant renin arterioles.** Every distant renin arteriole (more than 20 µm from every glomerulus, at least three renin cells) of treated and untreated patients with IgA nephropathy and of living donors among the 144 participants with a PAS-registered section, one block per group, ordered by renin cells (110-µm windows); title, renin cells in the wall. Tissue tiles as in Figure 1. IgAN, IgA nephropathy; JG, juxtaglomerular.

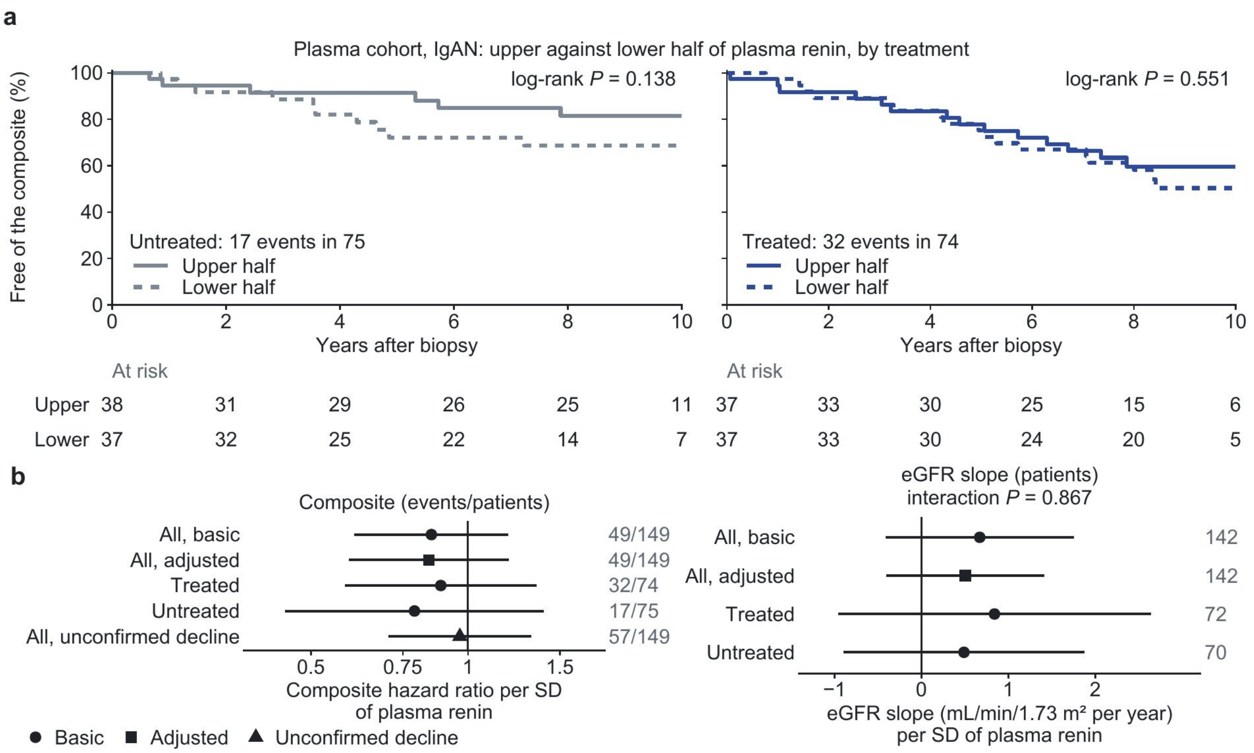


**Supplementary Figure S9. Plasma renin and kidney outcomes in the plasma cohort.** **a**, Kaplan–Meier curves of the composite outcome (eGFR decline of 40% or more from the eGFR at biopsy, confirmed at least 28 days later or, at the end of follow-up, without recovery to 80%, or eGFR below 15 mL/min/1.73 $m^2$ on two occasions) by plasma renin in the higher and lower half of each treatment group, in untreated and treated patients, with numbers at risk and log-rank *P*. **b**, Plasma renin per standard deviation: hazard ratio of the composite (logarithmic axis; basic and adjusted models, within each treatment group, and for the first decline without confirmation) and difference in eGFR slope (interaction *P*, plasma renin by treatment); events and patients at right. Points and lines, estimates and 95% confidence intervals. eGFR, estimated glomerular filtration rate; IgAN, IgA nephropathy; SD, standard deviation.
